# Semantic Segmentation of Sleep Events for High-Resolution Sleep Decoding

**DOI:** 10.1101/2025.09.25.25336636

**Authors:** Xiaoyu Bao, Man Li, Di Chen, Zijian Wang, Qiyun Huang, Zhenfu Wen, Yuanqing Li

## Abstract

Sleep is structured by brief, recurring EEG waveforms-such as slow waves, K-complexes, and spindles-that underpin sleep architecture and link to cognition, aging, and disease. Yet event-level analysis in sleep science remains constrained by reliance on labor-intensive manual annotation and the absence of automated, multi-event detection methods. Here, we present a unified, high-resolution framework for detecting multiple EEG-based sleep events continuously across the night. Integrating self-supervised and active learning to guide expert annotation, we constructed a cross-dataset, large-scale resource comprising 276,404 sleep events spanning seven physiologically and clinically relevant types. Leveraging this resource, we developed a sleep semantic segmentation model that decodes single-channel EEG into millisecond-level probability distributions for each event type. We demonstrated the versatility of the model across diverse applications in sleep science: real-time forecasting of imminent events to enable sleep interventions, automated sleep staging with state-of-the-art performance, and interpretable disease classification from whole-night EEG. By shifting sleep analysis from coarse staging to continuous, event-centric decoding, this study establishes a foundation for scalable, mechanistic, and clinically translatable sleep research.

## 1 Introduction

Sleep is a dynamic neurophysiological process composed of transient, recurring neural activities. Brief waveforms such as sleep spindles, K-complexes, and slow waves are increasingly recognized as the fundamental units of sleep architecture [1]. These sleep events, typically last from 0.5 to 3 seconds, not only delineate sleep stages but also reflect underlying cognitive processes, neurological functions, and psychiatric conditions [2]. In contrast to conventional sleep staging, which assigns fixed stage labels to 30-second epochs, sleep events provide a high-resolution, temporally precise view of sleep, grounded in quantifiable electrophysiological features [1].

A growing body of research has linked specific sleep events to diverse aspects of neurocognitive and clinical functions, including but not limit to sleep quality [3–5], cognitive abilities [6–8], aging [9–12], sex differences [13–16], and sleep-related disorders [17–20]. Specifically, sleep spindles and slow waves have been implicated in synaptic plasticity [21], memory consolidation [22], and aging-related shifts in brain dynamics [23]. K-complexes and micro-arousals reflect sleep stability and environmental responsiveness [24, 25], while less-characterized events such as sawtooth waves and vertex sharp waves are associated with REM-phase transitions and sensory processing [26, 27]. Even background EEG activity, often treated as a default sleep state, provides insights into cortical excitability and sleep continuity [28, 29]. Overall, these events form a continuous, multiscale signal landscape that reflects the structure and integrity of sleep.

While most prior studies have focused on individual sleep event types and their density, amplitude, or duration [30, 31], emerging evidence highlights the importance of temporal coordination and co-occurrence patterns across multiple events [32–34]. For example, slow waves facilitate an accurate temporal synchronization of sleep spindles, the quality of which is indicative of overnight memory retention [34]. Despite of the promising detection in examining multiple sleep events concurrently, progress remains constrained by the lack of automated, multi-event detection methods. Manual annotation of sleep events is time-consuming, expensive, requires domain expertise, and hard to scale, leading most studies to include only one or two event types, with limited sample size or recording duration [35, 36]. Although machine learning-based approaches are increasingly developed to automatically detect sleep events, they are often restricted to identifying a narrow subset of event types and lack generalizability across datasets [37–39].

To overcome these challenges, we proposed a hybrid strategy that combines selfsupervised learning (SSL) and active learning (AL) to guide efficient expert annotation (Figure 1). We used SSL to learn representations from unlabeled data, which facilitated initial model training and decrease the dataset size needed for initiating AL. The AL process iteratively identified the most informative segments for expert annotation, effectively accelerate the labeling process. Together, these approaches enabled us to compile a large-scale, cross-dataset resource comprising 276,404 sleep events spanning seven distinct types. Leveraging this resource, we developed a sleep semantic segmentation model (SSSM) that decodes single-channel EEG into a continuous, high-resolution probability distribution of sleep events at the sampling point level. This model enables a shift from coarse, stage-based analysis to event-centric decoding of sleep neurophysiology. We demonstrate the flexibility and effectiveness of the SSSM across three representative applications: (i) real-time forecasting of sleep events to support sleep interventions (ii) automatic sleep staging with state-of-the-art accuracy, and (iii) interpretable classification of sleep-related disorders from whole-night EEG.

**Fig. 1:**
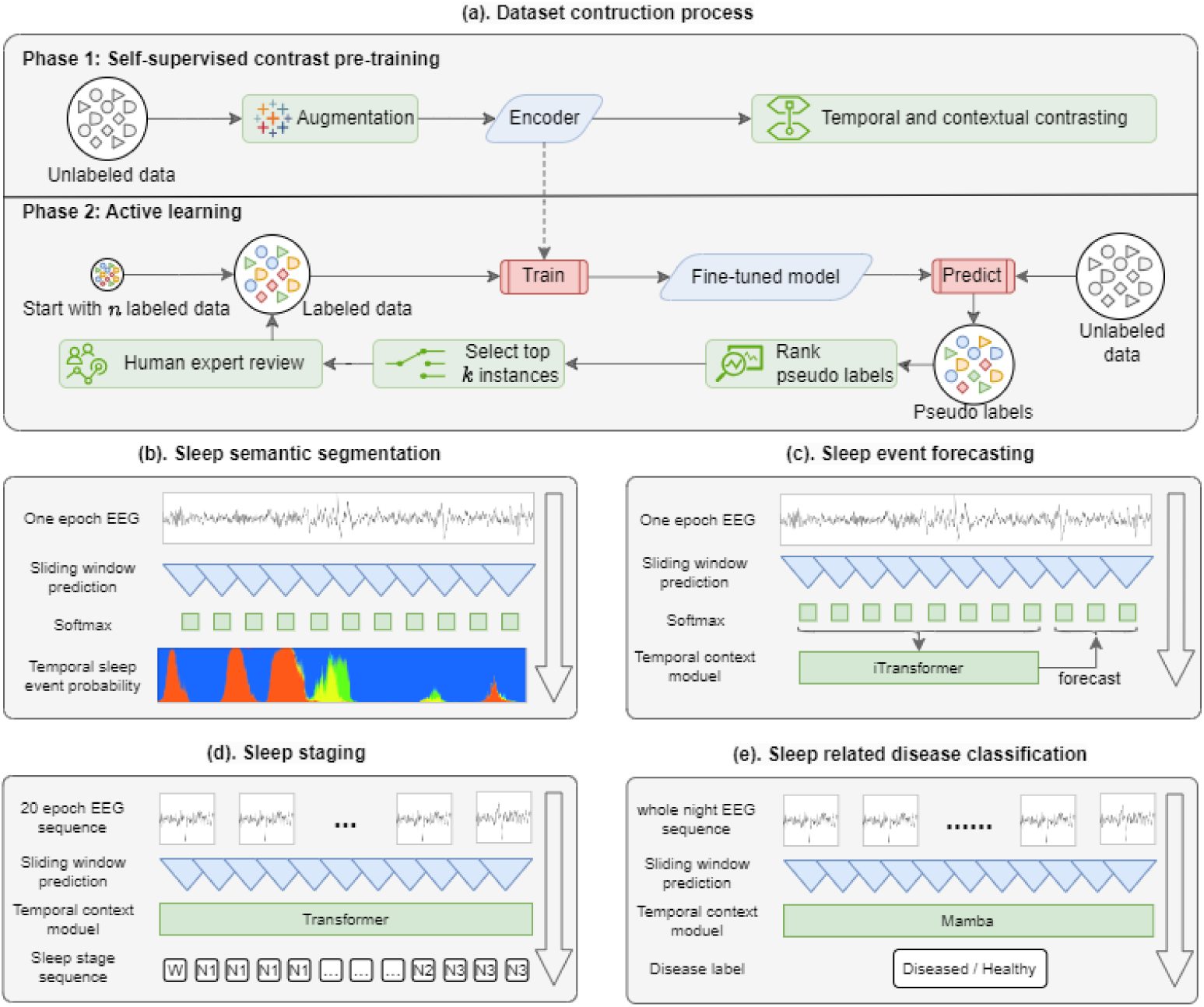
Methods overview. **(a).** Active learning process. Phase 1 involves a selfsupervised contrast learning approach. We employ completely unlabeled data and begin with an encoder that is initialized randomly. This encoder is then applied in Temporal and Contextual Contrasting self-supervised pretraining, resulting in a pretrained encoder. In Phase 2, this pre-trained encoder is integrated into the active learning cycle. **(b).** Configuration of the sleep semantic segmentation model. The base layer consists of single epoch EEG data (30 seconds). The next layer presents a sliding window of SSSM. Subsequently, multiple softmax modules form the third layer, leading to the output layer which provides sleep event probabilities. **(c).** Configuration of the sleep event forecasting model. In comparison to (b), an iTransformer acts as a temporal context module appended to the softmax layer, utilizing the previously predicted probabilities to forecast the masked likelihood of upcoming sleep events. **(d).** Configuration of the sleep staging model. Similar to (b), the initial layer is EEG signals, but this time comprising a sequence of 20 epochs. A Transformer encoder serving as a temporal context module is added to the sliding window SSSM, generating the sleep stage sequence. **(e).** Configuration of the sleep-related disease diagnosis model. In comparison to (c), the input layer is expanded to encompass a whole night EEG sequence, and the temporal context module could be Mamba. The resulting output layer provides disease labels.

## 2 Results

### 2.1 Construction of a Large-Scale Sleep Event Dataset

We built a large-scale, cross-dataset resource of sleep events by integrating selfsupervised learning (SSL) and active learning (AL) with expert annotation, enabling both efficient labeling and high-quality validation. The resulting dataset comprises 276,404 manually verified sleep events spanning seven types: background (BG), sleep spindle (SS), slow wave (SW), K-complex (KC), micro-arousal (MA), sawtooth wave (SAW), and vertex sharp wave (VSW). These sleep events were extracted from multiple publicly available sleep EEG datasets: Montreal Archive of Sleep Studies (MASS) [40], Sleep-EDF [41] , ISRUC [42] , and CAP [43]. These datasets collectively capture diverse demographics, clinical populations, and acquisition protocols, providing the breadth necessary for a robust and generalizable model foundation.

The annotation process began with an initial set derived from MASS, comprising 2,800 EEG segments (400 per event type) manually labeled by experts. This set was randomly split into a training set (*D*_init-train_) and a validation set (*D*_init-val_). We used *D*_init-train_ to train an event classification model which served as the starting point for AL, and used *D*_init-val_ to evaluate the classification performance on each AL iteration. To address the scarcity of labeled data (*D*_init-train_), we first applied SSL to pre-train the model on a large corpus of unlabeled EEG, yielding feature representations that were task-agnostic yet physiologically relevant. As shown in Figure 2a, cross-validation on *D*_init-train_ demonstrated that the SSL-based initialization substantially improved downstream classification performance compared to random initialization.

**Fig. 2:**
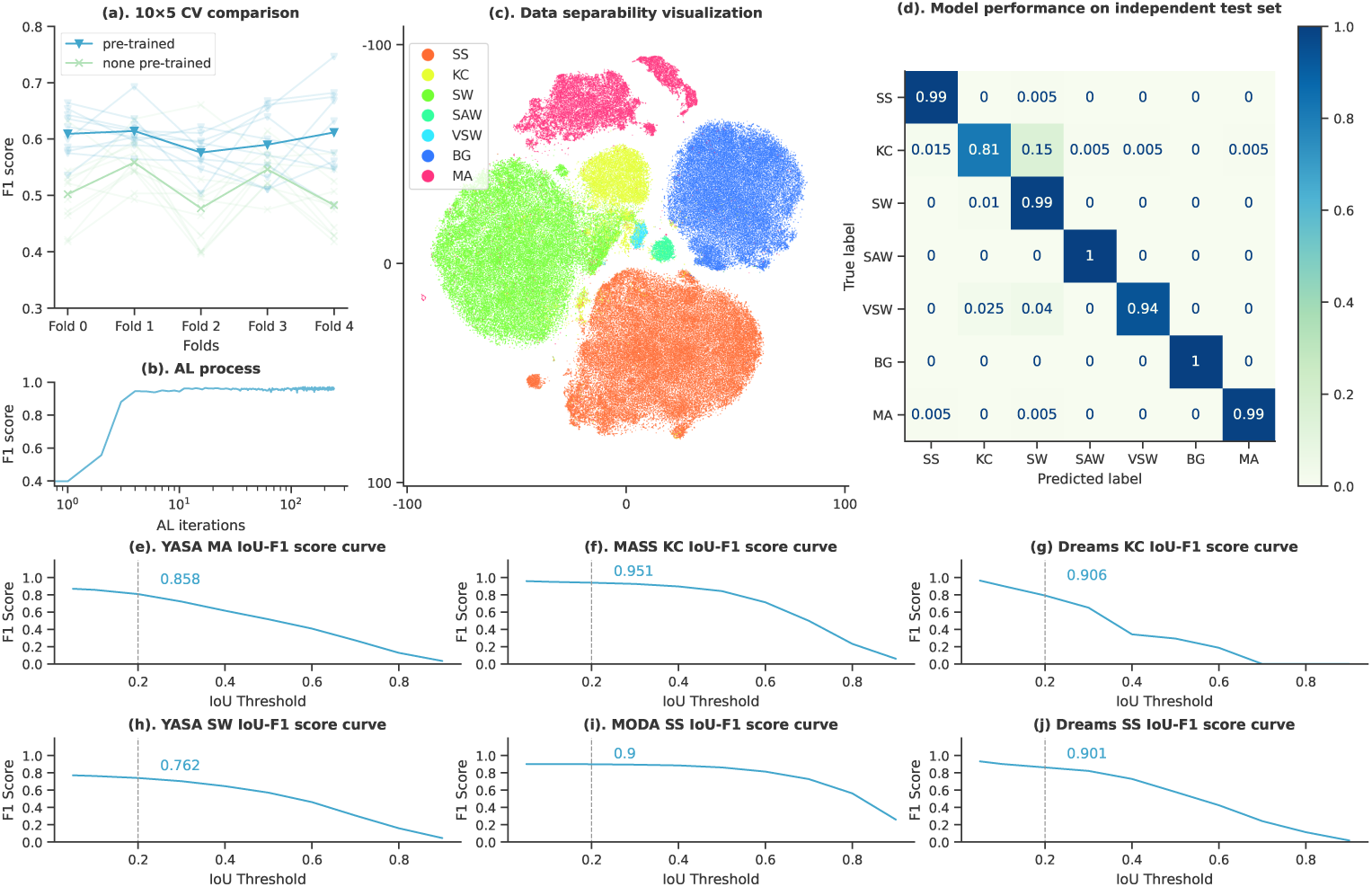
Dataset quality validation. **(a).** A 10×5 cross-validation assessment comparing a pre-trained model with a random initialized model The x-axis denotes the crossvalidation fold, while the y-axis represents the macro F1 score. Light blue illustrates the results of *F*_pre-trained_, and light green depicts the outcomes of *F*_random_. The lines connecting folds signify the same cross-validation test. Dark blue and dark green indicate the intra-group means. **(b).** The F1 score for sleep event classification improves as the number of AL iterations increases. **(c).** Data separability visualization. **(d).** Model performance on independent test set. **(e)-(j).** The F1-score-by-event in function of the IoU threshold. An overlap threshold of 0.2 (dotted vertical line) is used for evaluate the consensus of our dataset, as the F1 score of the cross point was annotated nearby. As the threshold increases (i.e. SSs must overlap with a higher percentage of the overall length of a compared event in order to be a match), the overall performance (F1) decreases.

The SSL-pretrained model was fine-tuned using *D*_init-train_ and then used as the backbone of an AL loop. In each AL iteration, unlabeled EEG segments were processed by the model, and segments with the highest prediction uncertainty (excluding BG) were prioritized for expert labeling. Newly labeled samples were incorporated into the training pool to retrain the classification model, and the selection process repeated, progressively improving its ability to select informative segments for annotation. This iterative process continued until all the data in the unlabeled pool were labeled on the MASS dataset (Figure 2b, evaluated on *D*_init-val_). The same AL strategy was subsequently applied to Sleep-EDF, ISRUC, and CAP. The AL cycles on these datasets were terminated once manual corrections fell below 10 per cycle, indicating that the model had successfully adapted to dataset-specific variability. In total, this pipeline produced 276,404 manually validated sleep events across seven types (Tabel 1), creating what is, to our knowledge, the largest rigorously validated, multi-event sleep dataset to date.

### 2.2 Validation of sleep event annotations

To evaluate the validity of our annotated sleep events, we performed a combination of qualitative and quantitative analyses across datasets and event types, assessing both the biological plausibility of the labels and the performance of the final classification model.

We first examined the separability of the annotated sleep events using the T-distributed Stochastic Neighbor Embedding (t-SNE). As shown in Figure 2c, the seven event types formed distinct, well-cohesive clusters with logical inter-cluster boundaries, reflecting their electrophysiological differences. We then evaluated the performance of the final classification model on *D*_init-val_, a manually labeled set that was never used for model training. At the conclusion of the AL process, the model achieved an average F1 score of 0.96 (Figure 2b), with the confusion matrix (Figure 2d) confirming strong discriminability across events and only minor overlap between SW and KC. Notably, the relatively lower separability between SW and KC mirrored their wellknown morphological similarity in human scoring practice [44]. These results indicates that the model-guided, expert-verified annotations are representative and internally consistent.

To benchmark against independent sources, we compared our annotations with labels from external expert scorers or state-of-the-art automatic detectors. Segment-level agreement was quantified using intersection-over-union (IoU) between predicted and reference event intervals, and IoU–F1 curves were constructed to visualize performance across thresholds. Following prior studies [35], we report F1 at IoU = 0.20 as the primary summary, balancing temporal tolerance with overlap strictness for short-lived, variably bounded sleep events. At this threshold, our KC annotations showed near-perfect concordance with the dual-expert MASS-SS2 labels (F1 = 0.951), and SS labels similarly aligned with MODA’s [35] crowdsourcing expert annotations (F1 = 0.900). For MA and SW—where large manually scored corpora are scarce—comparisons with YASA’s widely used detectors revealed good agreement (MA: F1 = 0.858; SW: F1 = 0.762).

To assess cross-dataset generalizability of our event detection model, we applied our trained model to the unseen Dreams dataset [45]. Our annotations remained highly consistent with expert labels for both KC (F1 = 0.906, Figure 3e) and SS (F1 = 0.901, Figure 3j).

**Fig. 3:**
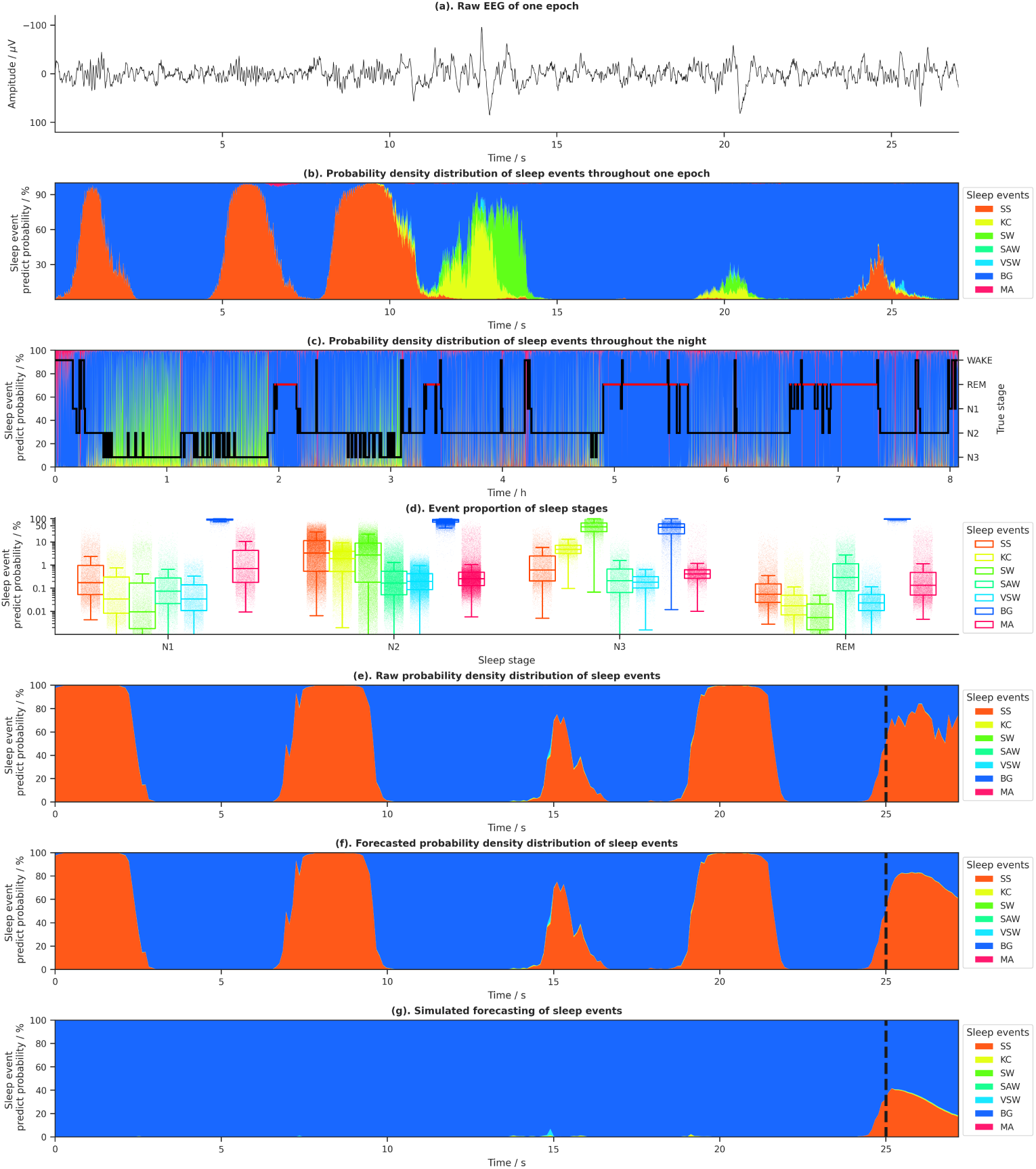
Performance of the sleep semantic segmentation model. **(a).** Raw EEG data of one N2 epoch (30 seconds). This epoch includes various BG, SS, KC, and SW events. **(b).** Probability density distribution of sleep events produced by the sleep semantic segmentation model, with each color corresponding to a different sleep event as shown in the legend besides. **(c).** Probability density distribution for throughout the night (left y-axis) alongside a ground truth hypnogram (right y-axis). **(d).** Event proportion of sleep stages. The probability density integral of each epoch was plotted as scatter points categorized by sleep stages, with an overlay of box plots. Each color represents a different sleep event, as indicated in the accompanying legend. **(e).** Raw probability density distribution of sleep events. **(f).** Forecast probability density distribution of sleep events. The predicted likelihood of sleep events is depicted as the right segment of black vertical line. **(g).** Simulated forecasting of sleep events. A portion of the probability density distribution, lacking SS events (though a minimal SS tail persists), 8 was simulated and entered into the sleep event forecasting model. The forecasted probability of sleep events is represented by the right section of the black vertical line.

Additionally, we performed an analysis of event occurrences on a per-subject basis using the MASS dataset. Differences in sleep event metrics related to age and gender were also examined. Detailed figures and statistical information can be found in the supplementary materials.

Collectively, the high cluster separability, strong held-out performance, IoU-based agreement with expert and benchmark annotations, and robustness on external datasets demonstrate that the SSL-AL pipeline produces event labels that are both biologically plausible and quantitatively reliable, providing a strong foundation for subsequent modeling and applications.

### 2.3 Sleep semantic segmentation model

Building on this rigorously validated resource, we developed the Sleep Semantic Segmentation Model (SSSM)—a neural architecture designed to translate single-channel EEG into temporally continuous, multi-event probability distributions. The term semantic segmentation is adapted from computer vision, where models assign a semantic label to every pixel of an image. Analogously, SSSM can assign a physiologically meaningful event label to every sampled time point of the EEG signal. Technically, SSSM extends the trained event classification backbone with a sliding-window strategy to output, for each time point, a vector of probabilities spanning all seven event types. Unlike conventional detectors that yield discrete, binary event markers, SSSM captures the graded onsets and offsets that characterize real neural events, offering a richer and more physiologically faithful representation of sleep dynamics.

As illustrated in Figure 3a–b, for representative raw EEG segments, SSSM generates smoothly varying, time-aligned probability curves for each event type, providing a richer view of neural dynamics than binary annotations. Applied to full-night recordings, these probability traces reveal structured patterns that qualitatively parallel traditional sleep staging (Figure 3c). Quantitative analysis of event probability distributions across sleep stages (Figure 3d) confirms close alignment with established physiology: in N1, micro-arousals (MA) follow background activity (BG) as the most prevalent; in N2, spindles (SS), K-complexes (KC), and vertex sharp waves (VSW) increase relative to N1; in N3, slow waves (SW) dominate while BGs diminish; and in REM, most event probabilities decrease, with BGs again prevailing. These stagespecific patterns are consistent with AASM guidelines regarding the occurrences of different events across sleep stages [46], underscoring the physiological fidelity of the model’s outputs.

Beyond accuracy, SSSM is computationally efficient: processing one hour of 100 Hz EEG requires only 52.9 ± 2.6 ms on a standard CPU and 6.44 ± 0.41 ms on a GPU, enabling real-time deployment in clinical workflows and closed-loop experimental paradigms. This combination of physiological validity, temporal granularity, and computational speed positions SSSM as a versatile backbone for downstream analyses and interventions.

#### 2.3.1 Application 1: forecasting imminent sleep events

One of the strengths of the SSSM framework is its ability to generate continuous, high-resolution probability trajectories for multiple event types across the entire night. These trajectories do not just describe the past events—they also contain rich temporal dependencies that govern how sleep events unfold, enabling the forecasting of imminent events. Such event forecasting has important implications: first, it enables closed-loop interventions that require precise timing, such as spindle-targeted memory reactivation; second, it provides a quantitative means to probe the causal and sequential structure of sleep neurophysiology. In the past, such forecasting has been hindered by the scarcity of large, multi-event annotated datasets and the lack of high-resolution segmentation models capable of producing temporally resolved event streams. Here, the high temporal-resolution multi-event probability streams generated by SSSM enabled us to investigate event forecasting.

We formulated event forecasting as a probability sequence modeling task: given the preceding 25 s of multi-event probability streams from SSSM, predict the probability distribution of events in the next 2 s. A transformer-based temporal architecture (iTransformer) was constructed and evaluated on the MASS-C3 dataset. This model achieved a mean squared error of 0.015 ± 0.001 and a mean absolute error of 0.043 ± 0.002. When predicted probabilities were converted to discrete labels, the prediction performance remained high (accuracy = 0.890 ± 0.011). To illustrate context sensitivity of the event forecasting model, we simulated examples to show how event distributions of preceding 25 s impacted the probability of upcoming sleep spindles (Figures 3e-g). When multiple spindles presented in the preceding window, the model forecasted high probabilities of imminent spindles (Figure 3e-f). By contrast, when only a trailing spindle tail was present, the predicted spindle probabilities collapsed (Figure 3g). This showed that the forecasting model leverages subtle temporal cues, rather than merely extrapolating from gross event presence.

We next quantified these dependencies systematically by manipulating either the past occurrence count or temporal proximity of SS, SW, and KC (Methods). This analysis revealed that for SS, both past occurrences (r = 0.450, p *<* 0.001) and temporal proximity (r = 0.957, p *<* 0.001) were strongly positively correlated with future occurrence probability, confirming the clustered nature of spindles. In contrast, SW exhibited a robust inhibitory effect on MA, with both higher past occurrence counts (r = –0.957, p *<* 0.001) and closer temporal proximity (r = –0.449, p = 0.043) suppressing future MA, consistent with the arousal-suppressing role of deep sleep. KC showed no significant relationship between its occurrence count and future SS probability (r = 0.068, p = 0.282), but its temporal proximity strongly predicted spindle onset (r = 0.995, p *<* 0.005), suggesting that KC–SS coupling is governed more by precise timing than by frequency.

Together, these findings establish SSSM as a framework in translating continuous event-level probabilities into event forecasts. Beyond enabling potential real-time interventions, such as targeted stimulation, the approach also provides a principled method for dissecting the fine-scale temporal dependencies that underlie sleep neurophysiology.

#### 2.3.2 Application 2: automatic sleep staging

Following event-level forecasting, we next asked whether SSSM’s continuous, highresolution representations could be aggregated to classify broader brain states—most notably, the canonical sleep stages. This task serves as a stringent test of the model’s representational power at a coarser temporal scale while leveraging the same backbone used for event segmentation.

We first tested SSSM in a single-epoch setting, appending a simple linear classification layer to the backbone. Using single-channel, 30-second EEG segments, the single-epoch model achieved performance comparable to, and in some cases exceeding, dedicated state-of-the-art sleep staging algorithms (refer to Supplementary materials, Table S2). This indicates that SSSM’s event-level features are informative and generalize effectively to stage classification without the need for specific fine-tuning. To capture multi-epoch dependencies, we introduced a Transformer layer on top of SSSM (SSSM-T), enabling the model to learn the structured transitions between contiguous epochs—a well-established way to improve classification performance. Across benchmark datasets, SSSM-T matched or outperformed state-of-the-art methods: in SHHS, it led in multiple metrics; in Sleep-EDF, it achieved top results in wake (W), N3, overall accuracy, and macro-F1; and in ISRUC-Sleep, it maintained the highest macro-F1 among all compared methods (refer to Supplementary materials, Table S3).

Considering the large variability across clinical sites, a model that is generalizable across different setting will be more useful and has higher potential for real-world deployment. We trained and evaluated the model using large-scale sleep EEG datasets from geographically dispersed clinical sites, spanning several decades and covering a wide range of demographics and patient groups (Table 2). Across 14 datasets, this model performed sleep staging with mean F1 ± STD (in parenthesis shown when weighted by number of test records) of 87.90±6.48 (88.70±7.96) for stage Wake, 43.96±5.04 (42.54±5.32) for stage N1, 81.70±1.67 (85.87±3.27) for N2, 77.18±5.26 (76.51±6.78) for stage N3, and 84.94±3.53 (85.78±3.02) for stage REM. The overall F1 performance can be summarized as 75.39 ± 3.58 (75.88 ± 4.09) ranging from a minimum of 70.38 (Challenge2018) to maximum 80.70 (SHHS) (refer to Supplementary materials, Table S4).

**Table 1:** The amount of annotated sleep events.

|  | BG | KC | MA | SAW | SW | SS | VSW | **Total** |
| --- | --- | --- | --- | --- | --- | --- | --- | --- |
| MASS | 50,770 | 15,551 | 26,388 | 2,598 | 70,293 | 77,387 | 1,601 | 244,588 |
| SleepEDF | 3,199 | 670 | 5,936 | 965 | 1,188 | 2,707 | 460 | 15,125 |
| ISRUC | 1,587 | 685 | 1,773 | 153 | 903 | 988 | 93 | 6,182 |
| CAP | 2,097 | 1,230 | 1,893 | 866 | 1,227 | 2,627 | 569 | 10,509 |
| **Total** | 57,653 | 18,136 | 35,990 | 4,582 | 73,611 | 83,709 | 2,723 | 276404 |

**Table 2:**
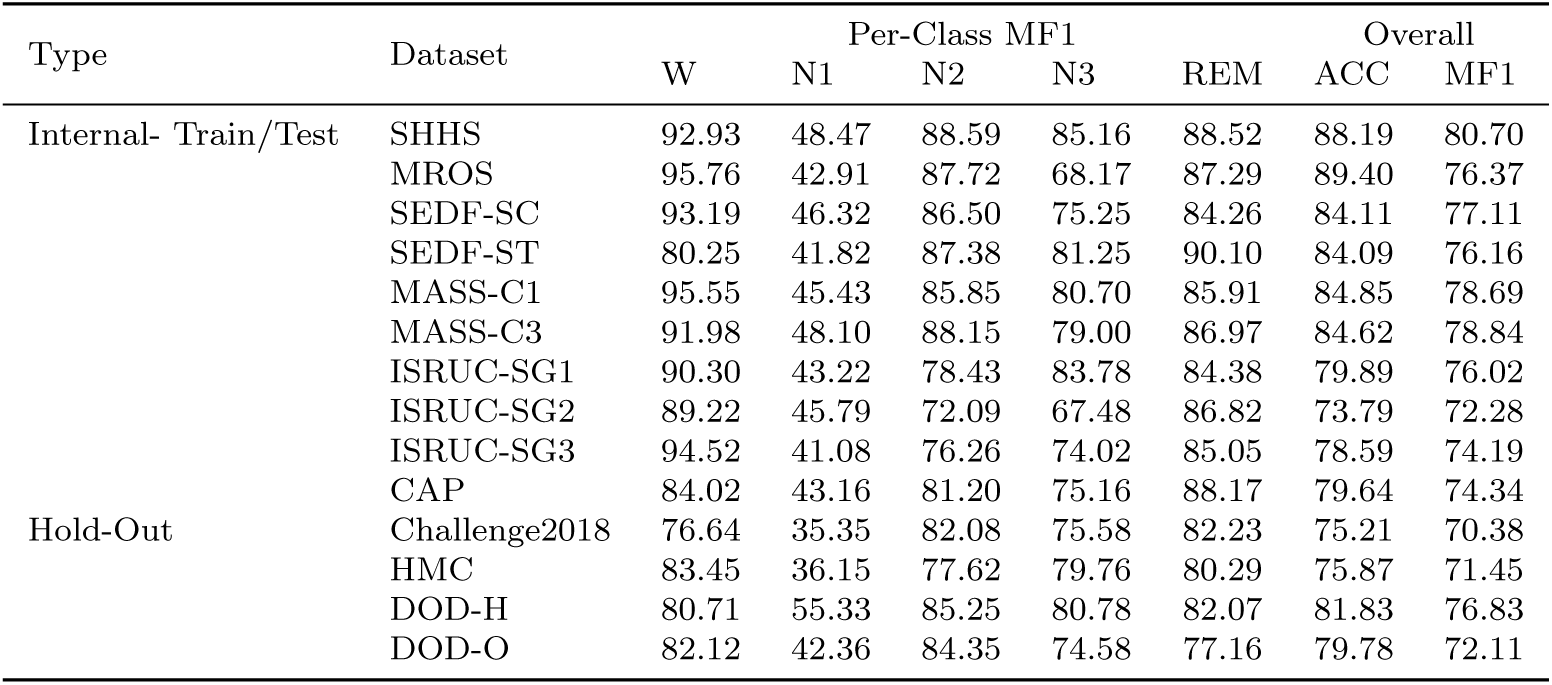
Cross-dataset Performance of the SSSM based Automatic Sleep Staging Model.

Together, these results show that SSSM’s event-centric, single-channel architecture scales seamlessly from fine-grained event detection (sleep semantic segmentation) to the clinically established but coarser task of sleep stage classification. Its realtime-capable design, combined with demonstrated cross-site robustness, makes it suitable for both laboratory and home-based monitoring, including large-scale clinical deployments.

#### 2.3.3 Application 3: sleep related disease diagnosis

Having shown that SSSM’s event-centric backbone can scale from fine-grained event mapping (sleep semantic segmentation) to coarse-grained sleep stage classification (Application 2), we next explored another challenging task: identifying pathological signatures from full-night EEG without hand-crafted features. This setting is particularly demanding because disease-relevant patterns are often sparse, brief, and embedded within hours of largely healthy background activity.

To address this, we integrated a sequence modeling module based on the Mamba architecture into SSSM, creating SSSM-Mamba. This addition enables the model to process entire overnight recordings while attending selectively to the most informative temporal segments. We evaluated SSSM-Mamba on three representative conditions: sleep apnea syndrome, REM behavior disorder (RBD), and depressive symptoms (Figures 4b, f and j). Across tasks, the model achieved strong classification performance, underscoring the transferability of event-centric representations to clinical diagnosis (refer to Supplementary materials, Table S5).

**Fig. 4:**
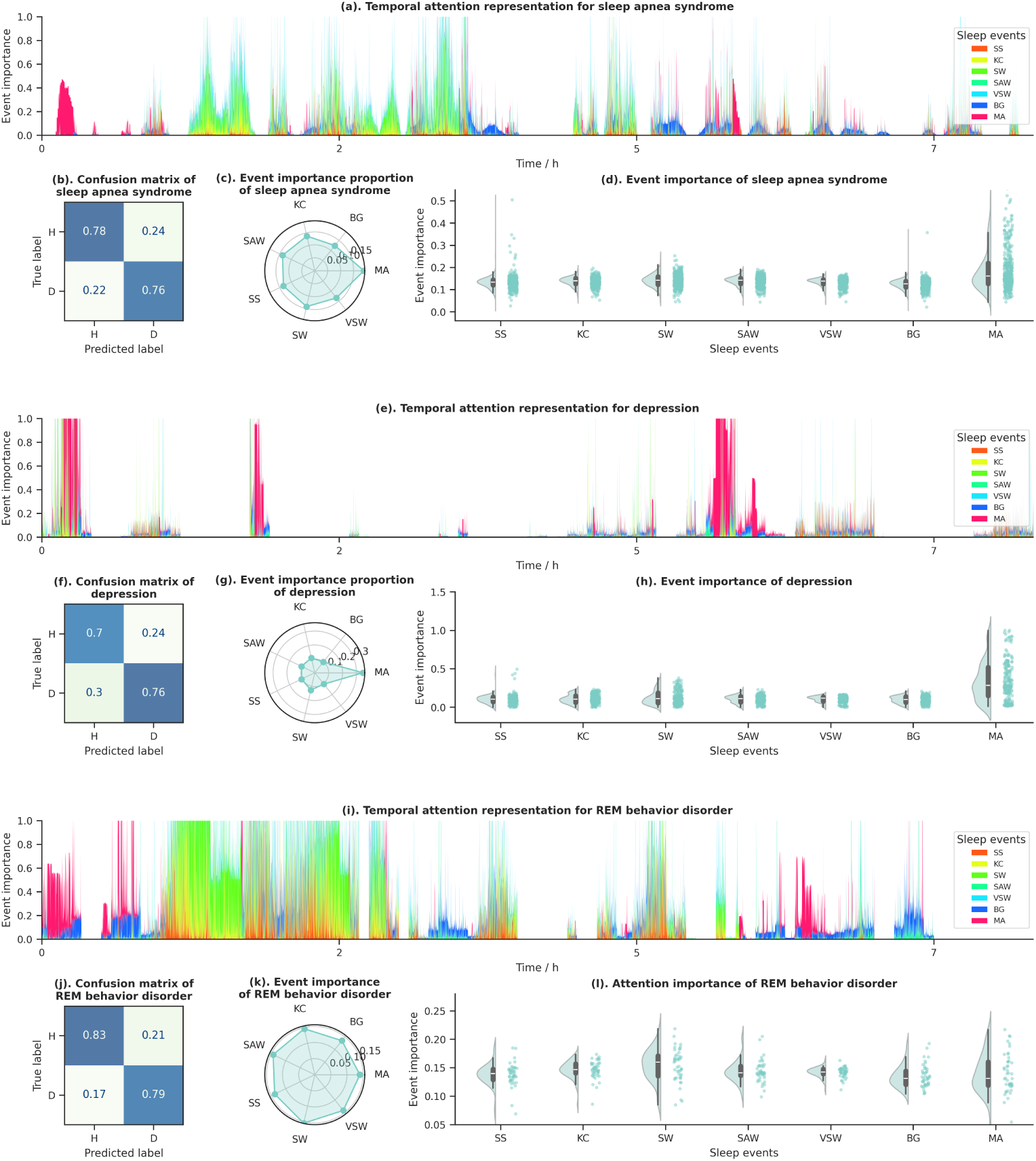
Model interpretability and exploration of disease-associated sleep events. **(a), (e) and (i).** Temporal attention representation for sleep apnea syndrome, depression, and REM behavior disorder, with different colors indicating sleep events as illustrated in the accompanying legend. **(b), (f) and (j).** Confusion matrix of sleep apnea syndrome, depression, and REM behavior disorder. **(c), (g) and (k).** Radar chart depicting attention proportions for sleep apnea syndrome, depression, and REM behavior disorder. **(d), (h) and (l).** Cloud-rain diagrams showcasing event importance for sleep apnea syndrome, depression, and REM behavior disorder.

A key advantage of SSSM-Mamba lies in its temporal attention mechanism, which makes the diagnostic process interpretable. As illustrated in Figures 4a, e, and i, attention maps reveal distinct, diagnosis-specific periods of interest. By aggregating these maps and quantifying the event composition within high-attention windows (Figures 4c, g and k), we identified condition-specific event contributions (Figures 4d, h, and l). For both sleep apnea and RBD, most event types contributed comparably to classification, with informative segments distributed broadly across the night. In contrast, depression classification relied disproportionately on micro-arousals (MA), which appeared in temporally clustered bursts—a pattern consistent with reported sleep disruption phenotypes in depression.

By combining continuous event-level representation with interpretable temporal attention, SSSM-Mamba transforms whole-night EEG from an opaque signal into a clinically interpretable map. This approach not only supports accurate diagnosis but also pinpoints the specific events and time windows driving model decisions, offering actionable insights for targeted interventions and mechanistic research.

## 3 Discussion

In this study, we present the first large-scale, cross-dataset resource of manually validated sleep events spanning seven distinct types, created through a hybrid selfsupervised learning (SSL) and active learning (AL) pipeline that efficiently directed expert effort toward the most informative samples. Leveraging this resource, we developed the Sleep Semantic Segmentation Model (SSSM), that transforms single-channel EEG into continuous, sampling-level probability distributions for multiple sleep events. This design moved beyond discrete, stage-based outputs toward a temporally resolved and physiologically faithful representation of sleep. We demonstrate the versatility of SSSM across three complementary applications: real-time forecasting of imminent events, automated sleep staging with state-of-the-art performance, and interpretable disease classification from whole-night EEG; highlighting its capacity to serve as a unified backbone for both mechanistic research and potential clinical applications. By uniting large-scale, multi-event annotation with a model capable of sampling-level temporal precision, this framework establishes a foundation for shifting sleep analysis from coarse epoch-level labels toward a richer, event-centric understanding of human sleep neurophysiology.

Sleep event analysis has long been constrained by the bottleneck of expert annotation. Manual labeling is labor-intensive, costly, and inherently narrow in scope, and existing studies rarely include more than one or two event types [35]. We addressed this challenge with a hybrid self-supervised learning (SSL) and active learning (AL) pipeline designed to maximize the value of each expert decision. SSL first distilled rich EEG representations from large unlabeled corpora, providing a stable foundation that requiring a smaller number of annotations for tuning the downstream event detection task. AL then strategically directed expert attention toward the most uncertain and informative samples, accelerating annotation while preserving accuracy. This process yielded 276,404 manually validated events spanning seven distinct types, drawn from four datasets with diverse demographics, recording environments, and clinical conditions. To our knowledge, this is the first dataset to combine such scale, diversity, and event breadth with rigorous manual verification. This dataset represents a valuable resource that can be used in future studies for different purposes, such as a developing and iteratively refining event-level computational models.

The validity of our sleep event dataset was supported by multiple, converging lines of evidence. Visualization of learned feature maps revealed clear separability among most event types, with the limited overlaps reflecting clinically recognized ambiguities in morphology or temporal context. For well-studied events—such as spindles, slow waves, K-complexes, and micro-arousals—our detection performance matched or surpassed existing public benchmarks, confirming the reliability of our annotations. For less characterized elements, including sawtooth waves, vertex sharp waves, and background activity, this work constitutes the first large-scale, rigorously validated corpus, filling a critical gap in the field. Beyond algorithmic metrics, the dataset reproduced established demographic trends—such as sex- and age-related differences in spindle density (Supplementary materials)—previously reported in large epidemiological cohorts [47–50], providing strong ecological validity. Together, these results demonstrate that the SSL–AL framework can generate data that are both broad in coverage and deep in quality, enabling applications that range from basic mechanistic research to clinically oriented translational studies.

Building on this resource, we developed the Sleep Semantic Segmentation Model (SSSM), trained across datasets that varied in reference electrodes, sampling rates, and acquisition protocols—demonstrating robustness to the heterogeneity that are typical in multi-center and real-world data. The SSSM operates with single-channel EEG, which maximizes portability and participant comfort, making it more suitable for continuous or longitudinal monitoring. Crucially, SSSM moves beyond conventional event detectors that produce discrete onset–offset markers, instead generating temporally continuous probability trajectories for each event type. This representation captures the inherently graded nature of sleep events—reflecting gradual transitions in amplitude and morphology—thereby providing a richer, more physiologically faithful depiction of the sleep landscape. This continuous mapping framework positions SSSM as a flexible backbone for diverse downstream applications, from mechanistic studies of event dynamics to real-time clinical interventions.

The ability to forecast imminent sleep events extends the utility of SSSM beyond descriptive analysis to predictive applications. Event forecasting has major implications for closed-loop interventions, such as slow-wave enhancement [51] or targeted memory reactivation [2] which require millisecond-level precision in event timing. Leveraging the continuous temporal probability trajectories output by SSSM, we trained a forecasting module to predict the occurrence of each event type up to two seconds in advance, using the preceding EEG activity. This approach achieved high predictive accuracy while faithfully capturing established physiological dependencies, such as the temporal clustering of spindles and the suppressive influence of slow waves on micro-arousals. Importantly, it also uncovered novel temporal associations, including the predictive role of K-complex onset for subsequent spindle initiation, suggesting that forecasting models may reveal previously overlooked mechanisms of event coupling. By moving from retrospective detection to prospective prediction, this framework creates new opportunities for precision-targeted interventions and causal testing of sleep-dependent processes.

Automatic sleep staging remains one of the most widely used EEG-based analyses, and our results show that SSSM’s learned representations perform competitively with or exceed state-of-the-art methods. In single-epoch evaluations, SSSM performed competitively with or exceed the accuracy of state-of-the-art staging algorithms [52, 53], suggesting that the generated continuous, multi-event probability landscapes are informative and effective for sleep staging. Extending to multi-epoch modeling with a transformer architecture (SSSM-T) improved performance further by capturing temporal dependencies between sleep stages. For large-scale clinical deployment, we developed a model trained on more than 10,000 PSGs from geographically and demographically diverse cohorts. Without fine-tuning or re-training, the model could be generalized across sites with high classification performance, demonstrating its potential for real-world applications. Importantly, in addition to stage labels, SSSM outputs the full probability distributions of multiple events, adding an extra analytical layer beyond conventional staging models.

SSSM also supports disease classification from full-night single-channel EEG, a task complicated by the fact that pathological signatures are often sparse, temporally fragmented, and embedded within otherwise normal activity. By integrating a Mamba sequence model with SSSM’s continuous event probability outputs, we developed SSSM-Mamba, which achieved high diagnostic accuracy across conditions including sleep apnea, REM behavior disorder, and depressive symptoms. Crucially, the temporal attention mechanisms of SSSM-Mamba yield interpretable links between model decisions and specific event patterns. For example, the model highlighted microarousals as dominant predictors in apnea and depression, and slow waves as critical indicators in REM behavior disorder. This event-level interpretability not only bridges the gap between black-box classification model and clinical trust but also offers mechanistic insight into how specific neural events contribute to disease phenotypes. By coupling high-resolution event decoding with transparent decision-making, this framework moves toward diagnostic tools that are both precise and explainable, enabling the potential for clinician–model collaboration in sleep medicine.

These applications demonstrate that continuous, event-level probability mapping is not merely a methodological upgrade but a reframing of how sleep neurophysiology can be represented and interrogated. By shifting from coarse, stage-based snapshots to a temporally resolved and physiologically grounded depiction of overlapping events, our framework allows questions to be asked at scales—both in time and population size—that were previously inaccessible. Scientifically, this enables systematic quantification of event co-occurrence, coupling dynamics, and inter-individual variability with a granularity that supports mechanistic hypotheses about memory consolidation, sensory processing, and neural regulation during sleep. It also opens the possibility of tracing developmental trajectories, mapping age- or sex-related changes, and identifying early deviations that precede clinical symptoms. Clinically, the framework enables personalized sleep phenotyping, where subtle changes in event structure over days or weeks could signal treatment response, cognitive decline, or emerging pathology. By conceptualizing sleep as a dynamic landscape of interacting neural events, this event-centric approach bridges macro-scale EEG markers with micro-scale neuronal circuit mechanisms, offering a natural interface for integration with other modalities such as fMRI and intracranial recordings.

Several limitations of this study should be considered. First, while the single-channel design offers clear advantages in terms of comfort, portability, and scalability, it inherently limits spatial analyses of event propagation and cortical topography. Extending SSSM to multi-channel configurations could yield richer mechanistic insights, particularly for investigating hemispheric asymmetries, regional differences, or localized pathology. Second, the curated event dataset was assembled from publicly available cohorts, which may underrepresent certain clinical populations, cultural contexts, or geographic regions. Incorporating data from wearable EEG systems and expanding coverage to broader age ranges could improve generalizability and applicability across diverse populations. Third, event definitions followed current AASM and field standards, but these taxonomies are not static. Future work may refine or expand event categories—potentially incorporating micro-events or novel elements revealed by high-density EEG—to capture a more complete picture of sleep neurophysiology. Fourth, for the disease diagnosis application, the sample sizes for each condition were relatively small. Larger, multi-site studies will be essential to validate diagnostic generalizability, assess robustness across heterogeneous clinical settings, and establish clinical utility.

In summary, this study presents the first large-scale, multi-event sleep dataset annotated through a hybrid SSL–AL strategy, and the first model to decode continuous, multi-event probability distributions from EEG at sampling-level resolution. By validating SSSM across diverse datasets and demonstrating its utility in staging, forecasting, and disease diagnosis, we establish it as both a scientific tool and a clinically relevant framework. Its flexibility and interpretability position it to drive advances in cognitive neuroscience, sleep medicine, and closed-loop intervention design. Looking forward, the combination of event-level precision, scalable architecture, and crossdomain adaptability may mark a shift from coarse, stage-based views of sleep toward a richer, temporally resolved understanding of human neurophysiology.

## 4 Methods

### 4.1 Construction of a large-scale sleep event dataset

#### 4.1.1 Data sources

The data employed for the pre-training of the backbone originated from the Montreal Archive of Sleep Studies (MASS) [40]. Subsequently, AL was applied to the whole MASS dataset via the pre-trained backbone. This choice was justified by MASS’s unique advantages for sleep event modeling: it is a large-scale, open-access polysomnography (PSG) database containing 200 complete overnight recordings from 200 participants (97 men, 103 women; age range: 18–76 years, mean: 38.3 years), covering diverse age groups and sleep phenotypes. Importantly, MASS includes human expert annotations for multiple sleep events (e.g., SS, KC), enabling the comparison of SSL-AL produced labels and human-annotated labels. To improve cross-dataset generalization, taking into account variations in recording protocols, demographics, and clinical populations, AL was later applied to three public sleep datasets: Sleep-EDF, ISRUC, and CAP. Notably, Sleep-EDF encompasses elderly participants, whereas CAP includes clinical populations with REM behavior disorder; further details of these public sleep datasets can be found in the supplementary materials.

#### 4.1.2 EEG preprocessing

To ensure the signal-to-noise ratio (SNR) and physiological relevance of the EEG data while unifying the data formats for subsequent model input, a standardized preprocessing workflow was implemented for the raw EEG signals. All preprocessing steps were implemented in Python via the mne library (version 2024.0 [54]), a tool widely validated in sleep EEG research for its consistency and adherence to neurophysiological data processing best practices. The specific steps, which are designed to retain sleep event-related neural activity while eliminating interference, are detailed below:

First, raw EEG data (acquired from datasets including MASS, Sleep-EDF, ISRUC, and CAP; see Data Sources section) were downsampled to 100 Hz via linear interpolation. This sampling rate was selected for two key reasons: 1) it preserves the critical frequency details of target sleep events—such as sleep spindles (11–16 Hz), slow waves (0.5–2 Hz), and K-complexes (0.5–4 Hz)—without information loss; and 2) it balances temporal resolution and computational efficiency, avoiding redundant data processing for subsequent event decoding.

Following downsampling, a finite impulse response filter with a Hamming window was applied to isolate frequencies relevant to sleep-wake oscillations. Key filter parameters were optimized for sleep EEG characteristics: FIR was chosen to avoid nonlinear phase distortion, ensuring that the temporal synchronization of sleep events is not skewed—critical for accurate event onset/offset detection; filter order was set to 200 taps, determined via the mne.filter.create filter function to achieve a narrow transition band (0.1 Hz) between the passband and stopband, minimizing signal attenuation near cutoff frequencies; and the frequency range was set to 0.3 Hz (lower cutoff) to 45 Hz (upper cutoff). The 0.3 Hz lower limit eliminates low-frequency artifacts, whereas the 45 Hz upper limit filters out high-frequency electromagnetic noise, aligning with the effective frequency range of sleep-related neural activity (0.5–40 Hz) as defined in sleep physiology standards.

#### 4.1.3 Expert annotation protocol

To ensure the reliability and consistency of sleep event labels, we established a rigorous expert annotation protocol, including strict scorer qualification criteria and a purpose-built scoring interface.

##### Scorer Qualifications

Ten expert scorers were recruited whose backgrounds included professional certification, academic training, and practical experience. This diversity ensured robust coverage of potential edge cases and alignment with clinical/research standards. Certified Specialists (Scorers 1–6): These scorers hold valid certification from the Board of Registered Polysomnographic Technologists (BRPT), the global gold standard for polysomnography (PSG) technical expertise. To obtain BRPT certification, candidates must succeed in a challenging examination that encompasses sleep physiology, EEG signal analysis, and adherence to AASM scoring guidelines, thereby guaranteeing that these scorers maintain clinical-level annotation accuracy. Academic Trainers (Scorers 7–8): Graduates of the Advanced Sleep Medicine Program at Southern Medical University Sleep Medicine Center (a leading institution for sleep research in China), these scorers have *>*3 years of hands-on experience annotating sleep events in both healthy and clinical populations. Their academic background ensures familiarity with cutting-edge sleep event characterization, complementing the clinical focus of BRPT-certified scorers. Experienced Practitioners (Scorers 9–10): With *>*2 years of dedicated sleep EEG annotation experience, these scorers have processed over 500 hours of PSG data across datasets prior to this study. Their expertise in handling dataset-specific variability helps minimize annotation bias across cross-dataset samples.

##### Collaborative Calibration

Before formal annotation, all ten scorers engaged in a collaborative calibration process. Each scorer independently annotated a shared training set consisting of three entire night EEG epochs, which included all seven target sleep events. Any discrepancies were resolved during group discussions led by a BRPT-certified scorer, with the AASM Manual for the scoring of sleep and associated events [46] serving as the definitive reference for classifying sleep events. Detailed information on the Guidelines for Annotating Sleep Events is available in the supplementary materials.

##### Scoring Interface Design and Operational Details

The annotation interface was configured via Label Studio [55], an open-source web-based platform specifically adapted for annotating time-series data, featuring design modifications suited to the specific requirements of sleep EEG event labeling. The raw EEG signals were segmented into 30-second epochs (consistent with the AASM sleep staging standards) and displayed one epoch at a time. This design minimized visual overload and ensured that scorers focused on fine-grained event details without distraction from adjacent epochs. The amplitude range is set to ±100 *µV*. The primary annotation took place on the C3 channel. Additional channels, such as Fpz and O2, were also shown for reference purposes, enabling scorers to crosscheck events such as the central dominance of a spindle and to eliminate artifacts specific to a channel, such as eye movement noise in Fpz. Scorers identified the beginning and end of sleep events through clickand-drag actions, with real-time feedback provided on the event’s duration (such as a spindle needing to last between 0.5 and 1.5 seconds according to the AASM guidelines). Notably, during the AL cycle, the interface was preloaded with a predetermined number of labels for experts to verify or amend, aiming to minimize annotation errors and expedite the annotation process. All annotations were timestamped with 100 Hz precision, aligned with the preprocessed EEG sampling rate, and associated with the original C3 channel data. This process ensured that the final annotated dataset used for SSSM training and validation was temporally precise and adhered to clinical and research standards.

#### 4.1.4 SSL-AL pipeline for sleep event annotation

To address the primary issue of expensive and low-efficiency manual annotation in sleep event labeling (compounded by the need to account for 7 event types over various datasets), we developed a hybrid SSL-AL pipeline. This methodology uses SSL to derive significant features from unlabeled data and employs AL to select high-information samples for expert annotation, maintaining an equilibrium between annotation quality, efficiency, and generalizability across datasets. The pipeline (depicted in Figure 1) is organized into two consecutive phases, with thorough validation conducted at each stage.

##### Phase 1: SSL Pre-Training for Feature Extraction

The goal of Phase 1 was to learn task-agnostic but physiologically relevant EEG representations from unlabeled data, reducing the number of labeled samples needed for subsequent initialization of AL. This phase was built on validated SSL methods for time-series data, with design choices tailored to sleep EEG characteristics. We employed a 3-block convolutional encoder [56], a configuration that has demonstrated efficacy for EEG time-series classification, as it effectively captures local temporal patterns. The encoder’s input was set to 1×300 (1 channel, 3 seconds of 100 Hz EEG), matching the preprocessed EEG segment size (see EEG Preprocessing section) and ensuring alignment with the downstream SSSM input. On the basis of the work of [57], which verified this SSL approach via EEG data, we applied contrastive learning to train the encoder. For each unlabeled EEG segment, we generated positive samples by applying small, physiologically plausible perturbations to preserve sleep event morphology. Negative samples were drawn from nonoverlapping EEG segments to ensure that the encoder learned to distinguish distinct neural patterns. We minimize contrastive loss, which maximizes the similarity between a segment and its positive samples while minimizing the similarity with negative samples. This forced the encoder to extract features that reflect sleep event physiology (e.g., frequency, amplitude) rather than noise. The encoder was pretrained on all unlabeled EEG segments from the MASS dataset (see Data sources section). For detailed implementation information, refer to [57].

##### Phase 2: AL Cycle for Targeted Annotation

Phase 2 integrates the SSL-pretrained encoder into an AL loop to boost the annotation process. To kickstart the AL loop, we constructed an independent initial dataset from the MASS dataset and randomly split it into two parts: *D*_init-train_ and *D*_init-val_, each including 1,400 samples with 200 samples per event type. *D*_init-train_ was used to fine-tune the pretrained encoder into a preliminary event classification model. *D*_init-val_ serves as a fixed validation benchmark to evaluate model performance in each AL iteration. Each AL cycle followed three steps and was repeated until the unlabeled data in the MASS were exhausted. Step 1, Pseudolabeling: The fine-tuned model (from the previous cycle) was applied to all unlabeled segments (initially from MASS) to generate pseudolabels and classwise probabilities (*p_k_* is the probability for the *k*-th event type). Step 2, Uncertainty Sampling: We rank the pseudolabeled samples by classification entropy (Eq. 1), a metric of model uncertainty [58, 59]:

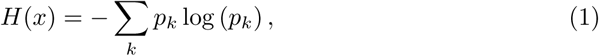

High entropy, indicated by values such as *H*(*x*) *>* 0.8, suggests that the model exhibits uncertainty concerning the event type of the sample; consequently, these particular samples are prioritized for annotation. For the BG, which accounts for a substantial proportion of sleep EEGs, per dataset statistic, active annotation was skipped to avoid redundant effort; instead, BG labels were indirectly determined by correcting pseudolabels of other events (e.g., reclassifying ambiguous “low-amplitude VSW” as the BG). Step 3, Expert Annotation and Model Update: In each iteration, the top 1,000 samples with the highest entropy (*k* = 1000) were chosen for manual annotation, as this quantity was determined to optimize efficiency and performance on the basis of preliminary experiments. These newly annotated samples were incorporated into *D*_init-train_, and the model underwent further fine-tuning.

##### Cross-Dataset AL Adaptation

After fully annotating the MASS dataset with AL, we adapted the workflow for three other datasets (Sleep-EDF, ISRUC, and CAP) by employing a restricted AL approach. Initially, the model fine-tuned on MASS was used to classify unlabeled segments in these datasets, creating pseudolabels. The AL process continued only until the number of necessary manual corrections decreased to fewer than 10 per cycle, which indicated sufficient adaptation to the unique characteristics of each dataset. Ultimately, this SSL and AL methodology produced 276,404 manually confirmed sleep events (refer to the Construction of large-scale sleep event dataset section). The persistent use of the pretrained encoder maintained feature uniformity, whereas AL reduced redundant annotations, providing solid ground for SSSM training.

#### 4.1.5 Dataset validation

To ensure the reliability, internal consistency, and external generalizability of the largescale sleep event dataset (*D*), which is essential for training the SSSM, we developed a comprehensive validation framework. This framework incorporates evaluations of model performance (validating the effectiveness of the SSL-AL pipeline), data separability (verifying the distinctness of event-specific features), and external consistency (aligning with established benchmarks). The specifics of each validation step, along with their rationale and results, are outlined below:

##### Evaluating the SSL-AL Pipeline’s Efficacy

This step aimed to validate whether the SSL-pretrained encoder alongside the iterative AL loop effectively enhanced the annotation quality and model performance, thus directly indicating the dataset’s ability to facilitate robust event classification. To quantify the value of SSL in extracting meaningful EEG features (vs. random initialization), we conducted 10×5-fold cross-validation on *D*_init-train_. This rigorous cross-validation design was chosen to minimize bias from random dataset splits. For each of the 10 independent iterations, *D*_init-train_ was randomly partitioned into 5 equal subsets. In each fold, 4 subsets were used for training (either with the SSL-pretrained encoder or a randomly initialized encoder), and the remaining subset was used for validation. This process is repeated 5 times per iteration (each subset serves as validation once), yielding two sets of F1 scores: *F*_pre-trained_ ∈ *R^i×j^*, F1 scores for the SSL-pretrained encoder (10 iterations × 5 folds), and *F*_random_ ∈ R*^i×j^*, F1 scores for the randomly initialized encoder.

##### Tracking AL Loop Performance

To ensure that the AL loop consistently improved dataset quality, we monitored model performance on *D*_init-val_ that, which was never used for training. After each AL iteration, we fine-tune the model and compute its macro-F1 score on *D*_init-val_. This process yields a sequence of scores *F*_AL_ ∈ R*^m^* (where *m* is the number of AL iterations). *D*_init-val_ served as a fixed “gold standard” to avoid overfitting to the expanding training set.

##### Verifying Event Feature Separability

To confirm that the dataset *D* captures distinct, physiologically meaningful patterns for each event type, we analyzed the high-dimensional feature output by the final AL-refined model via t-distributed stochastic neighbor embedding (t-SNE) (Maaten and Hinton, 2008). t-SNE converts high-dimensional feature similarities into low-dimensional joint probabilities, enabling visualization of whether distinct events form cohesive clusters. Clear clustering indicates that the dataset contains event-specific signals (not noise). Input *D* into the final AL-refined model to extract 1920-dimensional features (from the model’s output layer). t-SNE is applied to reduce the features to 2 dimensions, and then, clusters are visualized via seaborn (Waskom, 2021).

##### Quantifying Classification Accuracy with a Confusion Matrix

The confusion matrix quantifies true positives, false positives, and false negatives for each event, revealing whether the dataset’s labels are consistent with model expectations. To identify potential ambiguities in the dataset, we computed a confusion matrix by testing the final model on *D*_init-val_.

##### IoU-F1 curve: Defining a valid threshold

To maintain consistency of the dataset *D* with established sleep event annotations and algorithms, external consistency was assessed via the intersection over union (IoU), a standard metric in object detection/segmentation that measures the overlap between our labels and reference labels [35, 60]. The IoU measures the overlap between our dataset’s event intervals (D) and reference intervals and is calculated as follows: 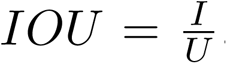,, where *I* denotes the area of intersection and *U* denotes the area of union. We generated IoU–F1 curves (F1 score vs. IoU threshold) to balance the localization accuracy (strict overlap) and detection completeness (tolerance for minor timing differences). In accordance with previous studies on sleep events [35, 60], we utilized an IoU threshold of 0.2, which effectively balances sensitivity to authentic events with resilience against minor discrepancies in annotation timing.

##### Validating specific event types

We assessed four primary event types (MA, SW, KC, and SS) against recognized references, as these events possess well-defined standards. For MA and SW, owing to the lack of publicly available datasets with manual MA/SW annotations, we utilized the extensively validated automatic detection tool yasa [61] to generate reference labels within the MASS dataset. For KC, the MASS-SS2 dataset [40], a specialized subset of MASS containing expert-annotated KCs (19 subjects, aged 23.6±3.7 years), was employed. For SS, validation was conducted against the MODA dataset [35], a crowdsourced dataset with expert-annotated spindles (containing 5,342 spindles from MASS). To ensure that dataset *D* facilitates model generalization, we applied the AL-refined model to the Dreams dataset [45], an independent dataset with expert-annotated SS and KC annotations that is not involved in any SSL/AL procedures. This finding demonstrates that dataset *D* effectively captures universal sleep event patterns rather than being limited to dataset-specific anomalies.

##### 4.2 Sleep semantic segmentation model design

The sleep semantic segmentation model (SSSM) was designed to provide continuous and high-resolution decoding of sleep events, particularly by translating single-channel EEG signals into millisecond-level probability distributions across seven different types of sleep events. The design emphasizes alignment with the physiological characteristics of sleep EEGs (such as the short-lived nature of most sleep events, which last between 0.5 and 3 s) and ensures its applicability in practical scenarios (for example, through real-time implementation and compatibility with constrained hardware environments). It consists of three main components: a backbone architecture optimized for time series data, a sliding window method to maintain temporal continuity, and a stringent assessment of computational efficiency.

#### 4.2.1 Backbone architecture

In Figure 1, the backbone of the SSSM is depicted as blue blocks, embodying a three-layer convolutional framework originally introduced by Wang et al. (2016) [56]. This design choice is underpinned by its proven efficacy in classifying time-series data. Unlike recurrent neural networks, which involve extra costs due to sequential processing, the convolutional approach focuses on detecting local temporal features such as the 11–16 Hz oscillatory pattern of sleep spindles (SS) or the biphasic waveform of K-complexes (KC), both of which are essential for the precise identification of sleep events. This approach also draws inspiration from Shelhamer et al. (2017) [62], who demonstrated that fully convolutional networks (FCNs) achieve semantic segmentation from “pixels to pixels” in computer vision; here, it is adapted to execute “time point-to-event probability” mapping in EEGs, a key function for real-time decoding of sleep events. The input setup of the backbone is characterized as “batch size × 1 × 300,” where “1” denotes the single-channel EEG and “300” represents 3 seconds of EEG data sampled at 100 Hz. This configuration is tailored to encapsulate the full duration of most sleep events while maintaining temporal resolution. Further details on the backbone parameters can be found in the Supplementary materials.

#### 4.2.2 Sliding window strategy

To address the limitation of fixed-epoch analysis, in which sleep events may span multiple 3-second intervals and thus be split or missed, a sliding window technique was integrated with the backbone, as shown in Figure 1b. This method converts 3-second chunks of EEG data into continuous event probability distributions, sampled at 100 Hz, encapsulating the concept of “sleep semantic segmentation.” The sliding window matches the input size of the backbone (3 seconds, 300 time points) and moves across the entire EEG record from night at 10-millisecond steps, corresponding to the EEG sampling rate, thereby ensuring temporal continuity in the output. A Softmax layer within the system generates a probability vector for each time point, ensuring that the total sum is 1 across the seven recognized sleep event categories. The event probability at any given EEG time point is assessed by the outputs from sliding windows centered on that time point.

#### 4.2.3 Model training

The SSSM training was tailored to emphasize robustness against variations in sleep events and ensure generalizability across different populations, which are essential for applying the model to real-world sleep analysis. Each phase was based on the characteristics of the extensive sleep event dataset *D* and the fundamental framework of the SSSM, as detailed below:

Initially, the 3-block convolutional backbone was initialized with weights obtained from the SSL pretraining step. This strategic decision was made to capitalize on EEG features relevant to sleep that were learned from unlabeled data. To prepare the samples, we standardized the input segments to accommodate the variable durations of sleep events in *D*, so they matched the 3-second (300 time points, 100 Hz) input dimension of the backbone. For every annotated event in *D*, we determined its midpoint (for example, a 1-second spindle occurring between 10 and 11 seconds in the EEG had a midpoint at 10.5 seconds) and extracted a ±1.5-second window centered on this midpoint. This chosen window size ensured comprehensive coverage of even the longest target events (3 s) and prevented the inclusion of unrelated adjacent activities. This standardization was crucial, as it aligned with the backbone’s receptive field, which is designed to capture 3 seconds of EEG data, thereby permitting the model to extract features consistently across all event types, irrespective of their initial duration. For events such as the BG, MA, and SW, which exceed 3 s, we randomly select nonoverlapping 3-second segments from the EEG intervals. Further details regarding the backbone parameters, including the size of the convolution kernel and the learning rate, can be found in the Supplementary materials.

#### 4.2.4 Computational Efficiency Validation

To ensure that the SSSM is applicable in practical scenarios (such as clinical sleep assessment and closed-loop interventions), it is essential that it runs efficiently on conventional hardware. To assess this efficiency, we tested the SSSM on two distinct systems: a CPU (11th Gen Intel® Core™ i7-11800H, 2.30 GHz) and a GPU (NVIDIA GeForce RTX 3060 Laptop GPU). The experimental dataset contained signals representing an hour of sampling at a frequency of 100 Hz, yielding a total of 360,000 data points. For each system configuration, seven separate trials were performed, each consisting of 10 execution cycles, totaling 70 iterations per platform. The mean and standard deviation of the processing time were determined across these iterations.

### 4.3 Downstream application configuration

#### 4.3.1 Forecasting imminent sleep events

Forecasting upcoming sleep events is an essential expansion of the sleep semantic segmentation model (SSSM)’s functionality, facilitating real-time, closed-loop interventions in sleep (such as spindle-targeted memory reactivation or enhancement of slow-wave activity) and mechanistic studies of the dynamics of sleep events. This endeavor utilizes the continuous, 100 Hz event probability outputs from the SSSM to capture temporal dependencies among events, with a process structured to balance physiological validity, model flexibility, and thorough validation.

##### Forecasting Model Design with iTransformer

Predicting upcoming sleep event probabilities from historical SSSM outputs is inherently a multivariate time series forecasting problem: the input is a sequence of 7-dimensional probability vectors (one vector per 10 ms time point, corresponding to the seven event types), and the output is a 7-dimensional vector of predicted probabilities for future time points. To address this issue, we chose the iTransformer [63], an enhanced version of the Transformer specifically designed for multivariate time series, as the temporal context module, which is appended to the SSSM’s Softmax layer (Figure 1c). The iTransformer was chosen over traditional Transformers or recurrent models (e.g., LSTMs) for two critical reasons. First, it rearranges the attention and feedforward layers across opposite dimensions. In contrast to conventional Transformers that integrate information across variables (event types) during the attention phase, the iTransformer processes each variable separately before aggregation, thereby preventing artificial cross-contamination of signals specific to each event. This aligns with the SSSM’s design, which treats each event as a distinct but cooccurring signal. Second, the iTransformer has demonstrated state-of-the-art performance on real-world multivariate datasets (e.g., physiological signals and environmental sensors), confirming its ability to capture the noisy, nonstationary temporal patterns inherent in sleep EEGs. For model configuration, we defined the input as the preceding 25 seconds of SSSM probability sequences (2,500 time points, 100 Hz) and the output as the subsequent 2 seconds of predicted probabilities (200 time points). This window size was optimized to balance two needs: (1) the 25-second input captures enough historical context to model event dependencies (e.g., the clustering of spindles every 5–10 s), and (2) the 2-second output encompasses the typical onset-to-peak duration of most sleep events (0.5–3 s), which is essential for interventions that need prior notification (e.g., initiating a stimulus 500 ms earlier than a forecasted SS). We trained and validated the iTransformer-SSSM pipeline exclusively on the MASS-C3 subset of MASS. MASS-C3 was selected for its demographic diversity (42.5 ± 18.9 years, 55% female) and high-quality EEG recordings, ensuring that the model learned generalizable temporal patterns rather than dataset-specific artifacts.

##### Validating Temporal Relationships between Key Event

To ensure that the forecasting model captured biologically meaningful temporal associations (not just statistical noise), we designed two targeted experiments focusing on four clinically relevant events: SS, SW, KC, and MA. These experiments quantified how two factors (past event occurrence count and temporal proximity) influence future event likelihood, with the results validated against known sleep physiology. Experiment 1, Influence of Past Event Occurrence Count: We synthesized datasets where the number of past occurrences of a target event (e.g., SS) was systematically varied (2, 3, 4, 5, or 6 occurrences) while controlling for the event interval (fixed at 5 seconds between consecutive events) and event tail (minimized to *<*0.1 seconds, ensuring no overlap between events). This control ensured that any changes in the predicted future probability were driven solely by the occurrence count, not the interval or residual activity. For each synthesized dataset, we input the past 25 seconds of probability sequences into the forecasting model and extract the predicted probability of the target event in the next 2 seconds. We then computed the Pearson correlation coefficient between the occurrence count and the predicted probability. Experiment 2, Influence of Temporal Proximity to Past Events: We synthesized datasets where the timing of a single past event was adjusted in 0.1-second increments (from 0 to 22 seconds before the forecast window), while again minimizing the event tail to avoid residual activity. This design tested how the recency of a past event affects future predictions.

#### 4.3.2 Automatic sleep staging

Automatic sleep staging assignment is a clinically foundational task that also serves as a critical test of the versatility of SSSM: it requires the translation of the model’s finegrained event-level representations into coarser, functionally meaningful brain states. Our approach leverages SSSM’s prelearned sleep event features to avoid redefining task-specific inputs while integrating temporal context (via Transformer encoders) to capture the sequential nature of sleep stages. Below is a detailed breakdown of the model’s architecture, underlying rationale, and thorough evaluation, all rooted in sleep physiology and aligned with demands for clinical applicability.

##### Single-Epoch Sleep Staging Model: Validating Event-Level Feature Utility

Initially, we created a single-epoch model to verify that the event-centric features of the SSSM can directly facilitate sleep staging. This model serves as a “baseline” to validate that the SSSM backbone captures EEG patterns relevant to stage classification without relying on a cross-epoch context. The input design of the model is intricately linked to the central architecture of SSSM: each 30-second EEG epoch, which serves as the standard unit for sleep staging, was divided into 10 distinct 3-second segments. These segments align with the 3-second input requirement of the SSSM convolutional backbone, involving a 100 Hz sampling rate and comprising 300 time points per segment. This partitioning ensures that we reuse the backbone’s pretrained event-extraction capabilities (e.g., detecting N2-specific spindles and N3-specific slow waves) without modifying its structure. Each 3-second segment was fed into the SSSM backbone, which outputs a feature vector encoding event-related patterns. These 10 feature vectors (one for each segment) were then integrated through average temporal pooling. This approach maintains the event distribution throughout the epoch (for example, an N3 epoch with 20 seconds of slow waves will exhibit greater N3 weights than an N2 epoch) while minimizing the impact of transient noise. The pooled vector was finally fed into a linear classifier, which mapped event features to one of five sleep stages. This minimalist design was purposefully employed: the utilization of a linear classifier allowed us to isolate the backbone features of the SSSM.

##### Multi-Epoch Sleep Staging Model (SSSM-T): Integrating the Temporal Context

Sleep staging is not just a per-epoch task: stages transition sequentially (e.g., N1 → N2 → N3, rarely N3 → Wake directly), and contextual information from adjacent epochs improves classification accuracy, especially for ambiguous stages such as N1 (which often lacks distinct events). To capture this sequentiality, we extended the single-epoch model with a Transformer encoder [64], creating the multi-epoch model SSSM-T (Figure 1d). The SSSM-T input was defined as a sequence of 20 consecutive epochs. Each of these 20 epochs was processed identically to the single-epoch model: partitioned into 10×3-second segments, fed into the SSSM backbone, and pooled into an epoch-level feature vector. This process yielded a sequence of 20 feature vectors (one per epoch), which was fed into a Transformer encoder with 8 attention heads and a model dimension of 512. Owing to the model’s efficiency and robustness, the SSSM backbone remained unchanged during Transformer training, whereas only the layers of the Transformer were subjected to fine-tuning. This choice preserves the backbone’s prelearned event features while optimizing the Transformer to model epoch-to-epoch transitions. The Transformer’s self-attention mechanism allows it to weigh the importance of adjacent epochs; for example, when classifying an ambiguous N1 epoch, it might prioritize features from the preceding Wake epoch and subsequent N2 epoch.

##### Clinical-Grade Evaluation: Ensuring Generalizability across Diverse Cohorts

To be of clinical utility, a sleep staging model should consistently perform across diverse populations that differ in demographics, recording methods, and clinical scenarios, thereby avoiding overfitting to limited datasets. We therefore conducted a large-scale evaluation using 14 publicly available polysomnographic (PSG) datasets (10,155 whole-night recordings total; further details on these datasets are provided in the Supplementary materials), spanning three decades of data collection, geographically dispersed clinical sites, and diverse groups (healthy adults, elderly individuals, patients with sleep apnea or REM behavior disorder). To mimic real-world clinical deployment, we split the 14 datasets into two groups with strict separation. Training/Internal Testing Group (10 datasets): This group included datasets such as MASS, Sleep-EDF, and SHHS. For each dataset, we used a per-subject split (75% training, 25% testing)—ensuring that no individual subject’s data appeared in both the training and testing sets (critical for avoiding overfitting, as a subject’s sleep patterns are consistent across epochs). All training data were used to refine SSSM-T into a clinical-ready variant (SSSM-B). Independent evaluation group (4 datasets): This dataset included the 2018 PhysioNet/CinC Challenge dataset, Haaglanden Medisch Centrum (HMC) dataset, and two Dream Open Datasets (DOD-H/O). Crucially, no data from these four datasets were used in any step of SSSM-B development. This design allowed us to assess “zero-shot” performance: how the model would perform when deployed to a new clinical site with unseen data. All datasets utilized are publicly available (with necessary approval for those with restricted access), ensuring transparency and reproducibility, which are essential requirements for implementing the model in clinical practice.

#### 4.3.3 Sleep related disease diagnosis

The use of sleep EEG to distinguish sleep-related disorders, such as REM behavior disorder (RBD), sleep apnea syndrome, and depression-related sleep disturbances, presents a unique technical challenge: it necessitates the examination of lengthy, continuous EEG recordings (spanning 8–10 hours overnight, or approximately ^~^10,000 3-second segments) while concurrently identifying two crucial feature types: (1) local pathological markers and (2) extended global patterns. Conventional sequence models face difficulties in balancing these requirements; Long Short-Term Memory networks (LSTMs) struggle with effectively capturing long-range dependencies, whereas standard Transformers [64], although proficient in sequence modeling, encounter *O*(*N* ^2^) computational complexity, rendering them unsuitable for full-night EEG analysis. To solve this issue, we combined the SSSM with Mamba [65], which is a structured state space sequence model optimized for lengthy sequences, forming a framework for disease diagnosis that achieves a balance among efficiency, feature detection, and clinical importance.

##### Mamba for Sleep Disease Diagnosis

Mamba’s architecture is specifically designed to address the shortcomings of traditional models, making it highly suitable for the overnight evaluation of EEGs, which is essential for diagnosing sleep disorders. It boasts features finely tuned to the needs of sleep-related diagnostics. Selective mechanism: Mamba differs from state space models that utilize fixed transition parameters by employing input-dependent parameters, enabling it to dynamically focus on clinically significant EEG segments (such as REM phases in patients with RBD and clusters of arousal induced by apnea during sleep apnea) while excluding irrelevant noise (such as stable background activity in healthy N2 sleep). Scan Module: By analyzing the EEG sequence in overlapping windows, the scan module can detect both local pathological markers (such as a 3-second microarousal) and long-range dependencies. This method mirrors the diagnostic approach that clinicians use for sleep disorders, which considers both transient events and the entire sleep pattern. Linear Computational Complexity: The hardware-optimized algorithm of Mamba ensures linear scalability with sequence length (*O*(*N* )), in contrast with transformers, which scale quadratically. This characteristic allows Mamba to efficiently process the complete set of 3-second EEG segments from an overnight study (up to 15,000 segments).

##### Mamba-SSSM Fusion Model Design

The disease diagnosis framework leverages the event-level feature extraction capabilities of the SSSM, incorporating a Mamba-based temporal context module to capture long-range EEG dynamics (Figure 1e). This approach allows us to utilize the SSSM’s preestablished sleep event features while introducing the capacity to associate these events with overarching disease patterns. The procedure is organized as follows. Step 1, Generating Disease-Relevant Token Features: Initially, we transform the complete overnight EEG into a sequence of event-level tokens via the SSSM backbone, maintaining the alignment with the core architecture of the model (as elaborated in “Sleep Semantic Segmentation Model Design”). For each 3-second EEG segment (corresponding to the backbone’s input size), the SSSM backbone derives a token that encapsulates event patterns. This token generation phase is vital: it translates raw EEG data into a succinct representation of sleep events, which serve as the biological substrates of disease-associated sleep phenotypes. Step 2, Mamba for Long-Sequence Classification: These tokens are subsequently input into a Mamba-based temporal context module, which learns to translate the sequence of event tokens into a disease label. More formally, the process is characterized as follows:

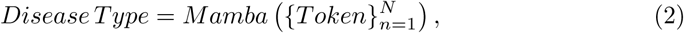

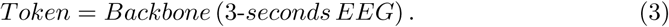

In this context, *N* refers to the number of 3-second intervals in an overnight recording, limited to a maximum of 15,000 (approximately 12.5 hours of EEG, which is more than sufficient for standard 8-10 hour sleep studies). For shorter recordings, such as a 6-hour EEG resulting in approximately 7,200 segments, we extend the token sequence with zeros to reach 15,000. This approach ensures a consistent input size across different subjects, thus preventing any bias toward longer recordings and facilitating batch training. Importantly, during Mamba training, the SSSM backbone remains unchanged, allowing only the Mamba module to be fine-tuned. This strategy maintains the backbone’s capacity to consistently extract event features while allowing the Mamba module to adapt and learn patterns specific to the disease.

##### Dataset Selection & Clinical Validation

To ensure that the model’s performance reflects real-world clinical utility, we evaluated it on three representative sleep disorders via two publicly available, clinically curated datasets: the CAP Sleep Database and the Sleep Heart Health Study (SHHS). Each dataset was selected for its ability to test the model’s ability to distinguish distinct pathological phenotypes, with strict sample selection criteria aligned with clinical standards. REM Behavior Disorder (RBD) –We used the CAP dataset, which includes 22 overnight recordings from patients with clinically diagnosed RBD and 16 recordings from age-matched healthy controls (those with no neurological or sleep disorders). Sleep Apnea Syndrome –We selected a subset of SHHS (a large community-based cohort) with clear clinical stratification. The apnea group included 262 subjects with an apnea–hypopnea index (AHI) *>* 50 events/hour (severe sleep apnea, per the AASM clinical criteria). Healthy control group: 215 subjects with both obstructive and central AHI *<* 5 events/hour (no clinically significant sleep apnea). This large sample size (477 total subjects) ensures that the model’s performance is not biased by small-sample variability, while the strict AHI thresholds align with how clinicians diagnose sleep apnea. Depression-Related Sleep Disturbance –We used another SHHS subset, where subjects were stratified by a validated “happy index” (a self-reported measure of mood correlated with depressive symptoms). Depression Group: Ninety-one subjects with a low happiness index (score *<* 3/5). Healthy control group: 130 subjects with a high happiness index (score *>* 4/5)—no self-reported mood disorders or sleep complaints. This stratification leverages SHHS’s comprehensive phenotypic data to ensure that the model is not just classifying arbitrary groups but also capturing sleep-related markers of a clinically relevant condition.

### 4.4 Construction of large-scale sleep event dataset

#### 4.4.1 Data sources

The data employed for the pre-training of the backbone originated from the Montreal Archive of Sleep Studies (MASS) [40]. Subsequently, AL was applied to the whole MASS dataset using the pre-trained backbone. This choice was justified by MASS’s unique advantages for sleep event modeling: it is a large-scale, open-access polysomnography (PSG) database containing 200 complete overnight recordings from 200 participants (97 men, 103 women; age range: 18–76 years, mean: 38.3 years), covering diverse age groups and sleep phenotypes. Importantly, MASS includes human expert annotations for multiple sleep events (e.g., SS, KC), enabling the comparation between SSL-AL produced label and human annotated label. In order to improve cross-dataset generalization, taking into account variations in recording protocols, demographics, and clinical populations, AL was later applied to three public sleep datasets: Sleep-EDF, ISRUC, and CAP. Notably, Sleep-EDF encompasses elderly participants, while CAP includes clinical populations with REM behavior disorder; further details of these public sleep datasets can be found in the supplementary materials.

#### 4.4.2 EEG preprocessing

To ensure the signal-to-noise ratio (SNR) and physiological relevance of EEG data, while unifying data formats for subsequent model input, a standardized preprocessing workflow was implemented for raw EEG signals. All preprocessing steps were implemented in Python using the mne library (version 2024.0 [54]), a tool widely validated in sleep EEG research for its consistency and adherence to neurophysiological data processing best practices. The specific steps, designed to retain sleep-event-related neural activity while eliminating interference, are detailed below:

First, raw EEG data (acquired from datasets including MASS, Sleep-EDF, ISRUC, and CAP; see Data Sources section) was downsampled to 100 Hz using linear interpolation. This sampling rate was selected for two key reasons: 1) it preserves the critical frequency details of target sleep events—such as sleep spindles (11–16 Hz), slow waves (0.5–2 Hz), and K-complexes (0.5–4 Hz)—without information loss; 2) it balances temporal resolution and computational efficiency, avoiding redundant data processing for subsequent event decoding.

Following downsampling, a finite impulse response filter with a Hamming window was applied to isolate frequencies relevant to sleep-wake oscillations. Key filter parameters were optimized for sleep EEG characteristics: FIR was chosen to avoid nonlinear phase distortion, ensuring the temporal synchronization of sleep events is not skewed—critical for accurate event onset/offset detection; Filter order set to 200 taps, determined via the mne.filter.create filter function to achieve a narrow transition band (0.1 Hz) between passband and stopband, minimizing signal attenuation near cutoff frequencies; Frequency range set to 0.3 Hz (lower cutoff) to 45 Hz (upper cutoff). The 0.3 Hz lower limit eliminates low-frequency artifacts, while the 45 Hz upper limit filters out high-frequency electromagnetic noise, aligning with the effective frequency range of sleep-related neural activity (0.5–40 Hz) as defined in sleep physiology standards.

#### 4.4.3 Expert annotation protocol

To ensure the reliability and consistency of sleep event labels, we established a rigorous expert annotation protocol, including strict scorer qualification criteria and a purpose-built scoring interface.

##### Scorer Qualifications

Ten expert scorers were recruited, with their backgrounds covered professional certification, academic training, and practical experience. This diversity ensured robust coverage of potential edge cases and alignment with clinical/research standards. Certified Specialists (Scorers 1–6): These scorers hold valid certification from The Board of Registered Polysomnographic Technologists (BRPT), the global gold standard for polysomnography (PSG) technical expertise. To obtain BRPT certification, candidates must succeed in a challenging examination that encompasses sleep physiology, EEG signal analysis, and adherence to AASM scoring guidelines, thereby guaranteeing that these scorers maintain clinical-level annotation accuracy. Academic Trainers (Scorers 7–8): Graduates of the Advanced Sleep Medicine Program at Southern Medical University Sleep Medicine Center (a leading institution for sleep research in China), these scorers have *>*3 years of hands-on experience annotating sleep events in both healthy and clinical populations. Their academic background ensures familiarity with cutting-edge sleep event characterization, complementing the clinical focus of BRPT-certified scorers. Experienced Practitioners (Scorers 9–10): With *>*2 years of dedicated sleep EEG annotation experience, these scorers have processed over 500 hours of PSG data across datasets prior to this study. Their expertise in handling dataset-specific variability helps minimize annotation bias across cross-dataset samples.

##### Collaborative Calibration

Before formal annotation, all ten scorers engaged in a collaborative calibration process. Each scorer independently annotated a shared training set consisting of three entire night EEG epochs, which included all seven target sleep events. Any discrepancies were resolved during group discussions led by a BRPT-certified scorer, with the AASM Manual for the scoring of sleep and associated events [46] serving as the definitive reference for classifying sleep events. Detailed information on the Guidelines for Annotating Sleep Events is available in the supplementary materials.

##### Scoring Interface Design and Operational Details

The annotation interface was configured using Label Studio [55], an open-source web-based platform specifically adapted for annotating time-series data, featuring design modifications suited to the specific requirements of sleep EEG event labeling. Raw EEG signals were segmented into 30-second epochs (consistent with AASM sleep staging standards) and displayed one epoch at a time. This design minimized visual overload and ensured scorers focused on fine-grained event details without distraction from adjacent epochs. Amplitude range set to ±100 *µV*. The primary annotation took place on the C3 channel. Additional channels, such as Fpz and O2, were also shown for reference purposes, enabling scorers to cross-check events like the central dominance of a spindle and to eliminate artifacts specific to a channel, such as eye movement noise in Fpz. Scorers identified the beginning and end of sleep events through click-and-drag actions, with real-time feedback provided on the event’s duration (such as a spindle needing to last between 0.5 and 1.5 seconds according to AASM guidelines). Notably, during the AL cycle, the interface was preloaded with a predetermined number of labels for experts to verify or amend, aiming to minimize annotation errors and expedite the annotation process. All annotations were timestamped with 100 Hz precision, aligning with the preprocessed EEG sampling rate, and associated with the original C3 channel data. This process ensured that the final annotated dataset used for SSSM training and validation was temporally precise and adhered to clinical and research standards.

#### 4.4.4 SSL-AL pipeline for sleep event annotation

To tackle the primary issue of expensive and low-efficiency manual annotation in sleep event labeling (compounded by the necessity to account for 7 event types over various datasets), we developed a hybrid SSL-AL pipeline. This methodology uses SSL to derive significant features from unlabeled data and employs AL to select high-information samples for expert annotation, maintaining an equilibrium between annotation quality, efficiency, and generalization across datasets. The pipeline (depicted in Figure 1) is organized into two consecutive phases, with thorough validation conducted at each stage.

##### Phase 1: SSL Pre-Training for Feature Extraction

The goal of Phase 1 was to learn task-agnostic but physiologically relevant EEG representations from unlabeled data, reducing the number of labeled samples needed for subsequent initialization of AL. This phase built on validated SSL methods for time-series data, with design choices tailored to sleep EEG characteristics. We employed a 3-block convolutional encoder [56], a configuration that has demonstrated efficacy for EEG time-series classification, as it effectively captures local temporal patterns. The encoder’s input was set to 1×300 (1 channel, 3 seconds of 100 Hz EEG), matching the preprocessed EEG segment size (see EEG Preprocessing section) and ensuring alignment with downstream SSSM input. Based on the work by [57], which verified this SSL approach using EEG data, we applied contrastive learning to train the encoder. For each unlabeled EEG segment, we generated positive samples by applying small, physiologically plausible perturbations to preserve sleep event morphology. Negative samples were drawn from non-overlapping EEG segments to ensure the encoder learns to distinguish distinct neural patterns. We minimized the contrastive loss, which maximizes similarity between a segment and its positive samples while minimizing similarity with negative samples. This forced the encoder to extract features that reflect sleep event physiology (e.g., frequency, amplitude) rather than noise. The encoder was pre-trained on all unlabeled EEG segments from the MASS dataset (see Data sources section). For detailed implementation information, refer to [57].

##### Phase 2: AL Cycle for Targeted Annotation

Phase 2 integrated the SSL-pre-trained encoder into an AL loop to boost annotation process. To kickstart the AL loop, we constructed an independent initial dataset from the MASS dataset and randomly split it into two: *D*_init-train_ and *D*_init-val_, each include 1,400 samples with 200 samples per event type. *D*_init-train_ was used to fine-tune the pre-trained encoder into a preliminary event classification model. *D*_init-val_ served as a fixed validation benchmark to evaluate model performance in each AL iteration. Each AL cycle followed three steps, repeated until the unlabeled data in the MASS is Exhausted. Step 1, Pseudo-Labeling: The fine-tuned model (from the previous cycle) was applied to all unlabeled segments (initially from MASS) to generate pseudo-labels and class-wise probabilities (*p_k_* is the probability for the *k*-th event type). Step 2, Uncertainty Sampling: We ranked pseudo-labeled samples by classification entropy (Eq. 1), a metric of model uncertainty [58, 59]:

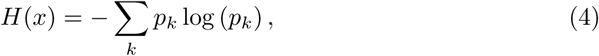

High entropy, indicated by values like *H*(*x*) *>* 0.8, suggests that the model exhibits uncertainty concerning the event type of the sample; consequently, these particular samples are prioritized for annotation. For BG, which accounts for a substantial proportion of sleep EEG, per dataset statistics, active annotation was skipped to avoid redundant effort; instead, BG labels were indirectly determined by correcting pseudolabels of other events (e.g., reclassifying ambiguous “low-amplitude VSW” as BG). Step 3, Expert Annotation and Model Update: In each iteration, the top 1,000 samples with the highest entropy (*k* = 1000) were chosen for manual annotation, as this quantity was determined to optimize efficiency and performance based on preliminary experiments. These newly annotated samples were incorporated into *D*_init-train_, and the model underwent further fine-tuning.

##### Cross-Dataset AL Adaptation

After fully annotating the MASS dataset with AL, we adapted the workflow for three other datasets (Sleep-EDF, ISRUC, CAP) by employing a restricted AL approach. Initially, the model fine-tuned on MASS was used to classify unlabeled segments in these datasets, creating pseudo-labels. The AL process continued only until the necessary manual corrections dropped to fewer than 10 per cycle, which indicated sufficient adaptation to the unique characteristics of each dataset. Ultimately, this SSL and AL methodology produced 276,404 manually confirmed sleep events (refer to the Construction of large-scale sleep event dataset section). The persistent use of the pre-trained encoder maintained feature uniformity, whereas AL reduced redundant annotation, providing a solid ground for SSSM training.

#### 4.4.5 Dataset validation

Dataset validation

To ensure the reliability, internal consistency, and external generalizability of the large-scale sleep event dataset (*D*), essential for training the SSSM, we developed a comprehensive validation framework. This framework incorporates evaluations of model performance (validating the effectiveness of the SSL-AL pipeline), data separability (verifying the distinctness of event-specific features), and external consistency (aligning with established benchmarks). The specifics of each validation step, along with their rationale and results, are outlined below:

##### Evaluating the SSL-AL Pipeline’s Efficacy

This step aimed to validate if the SSL-pre-trained encoder alongside the iterative AL loop effectively enhanced annotation quality and model performance, thus directly indicating the dataset’s capability to facilitate robust event classification. To quantify the value of SSL in extracting meaningful EEG features (vs. random initialization), we conducted 10×5-fold crossvalidation on *D*_init-train_. This rigorous cross-validation design was chosen to minimize bias from random dataset splits. For each of 10 independent iterations, *D*_init-train_ was randomly partitioned into 5 equal subsets. In each fold, 4 subsets were used for training (either with the SSL-pre-trained encoder or a randomly initialized encoder), and the remaining subset was used for validation. This process repeated 5 times per iteration (each subset serving as validation once), yielding two sets of F1 scores: *F*_pre-trained_ ∈ *R^i×j^*, F1 scores for the SSL-pre-trained encoder (10 iterations × 5 folds), and *F*_random_ ∈ R*^i×j^*, F1 scores for the randomly initialized encoder.

##### Tracking AL Loop Performance

To ensure the AL loop consistently improved dataset quality, we monitored model performance on *D*_init-val_ that was never used for training. After each AL iteration, we fine-tuned the model and computed its macro-F1 score on *D*_init-val_. This yielded a sequence of scores *F*_AL_ ∈ R*^m^* (where *m* is the number of AL iterations). *D*_init-val_ served as a fixed “gold standard” to avoid overfitting to the expanding training set.

##### Verifying Event Feature Separability

To confirm that the dataset *D* captures distinct, physiologically meaningful patterns for each event type, we analyzed the high-dimensional features output by the final AL-refined model using t-SNE (t-Distributed Stochastic Neighbor Embedding) (Maaten and Hinton, 2008). t-SNE converts high-dimensional feature similarities into low-dimensional joint probabilities, enabling visualization of whether distinct events form cohesive clusters. Clear clustering indicates the dataset contains event-specific signals (not noise). Input *D* into the final AL-refined model to extract 1920-dimensional features (from the model’s output layer). Apply t-SNE to reduce the features to 2 dimensions, then visualize clusters using seaborn (Waskom, 2021).

##### Quantifying Classification Accuracy with Confusion Matrix

The confusion matrix quantifies true positives, false positives, and false negatives for each event, revealing whether the dataset’s labels are consistent with model expectations. To identify potential ambiguities in the dataset, we computed a confusion matrix by testing the final model on *D*_init-val_.

##### IoU-F1 Curve: Defining a Valid Threshold

To maintain consistency of the dataset *D* with established sleep event annotations and algorithms, external consistency was assessed utilizing the Intersection over Union (IoU), a standard metric in object detection/segmentation that measures the overlap between our labels and reference labels [35, 60]. IoU measures the overlap between our dataset’s event intervals (D) and reference intervals, calculated as: 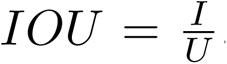, where, *I* donates the area of intersection, and *U* donates the area of union. We generated IoU-F1 curves (F1 score vs. IoU threshold) to balance localization accuracy (strict overlap) and detection completeness (tolerance for minor timing differences). In accordance with previous studies on sleep events [35, 60], we utilized an IoU threshold of 0.2, which effectively balances sensitivity to authentic events with resilience against minor discrepancies in annotation timing.

##### Validating Specific Event Types

We assessed four primary event types (MA, SW, KC, SS) against recognized references, as these events possess well-defined standards. For MA and SW: Due to the lack of publicly available datasets with manual MA/SW annotations, we utilized the extensively validated automatic detection tool, yasa [61], to generate reference labels within the MASS dataset. For KC: The MASS-SS2 dataset [40], a specialized subset of MASS containing expert-annotated KCs (19 subjects, aged 23.6±3.7 years), was employed. For SS: Validation was conducted against the MODA dataset [35], a crowdsourced dataset with expert-annotated spindles (containing 5,342 spindles from MASS). To ensure dataset *D* facilitates model generalization, we applied the AL-refined model to the Dreams dataset [45], an independent dataset with expert-annotated SS and KC annotations, not involved in any SSL/AL procedures. This demonstrates dataset *D* effectively captures universal sleep event patterns, rather than being limited to dataset-specific anomalies.

### 4.5 Sleep semantic segmentation model design

The Sleep Semantic Segmentation Model (SSSM) was designed to provide continuous and high-resolution decoding of sleep events, particularly by translating single-channel EEG signals into millisecond-level probability distributions across seven different types of sleep events. The design emphasizes alignment with the physiological characteristics of sleep EEG (such as the short-lived nature of most sleep events, which last between 0.5 to 3 seconds) and ensures its applicability in practical scenarios (for instance, through real-time implementation and compatibility with constrained hardware environments). It consists of three main components: a backbone architecture optimized for time-series data, a sliding window method to maintain temporal continuity, and stringent assessment of computational efficiency.

#### 4.5.1 Backbone architecture

In Figure 1, the backbone of the SSSM is depicted as blue blocks, embodying a three-layer convolutional framework originally introduced by Wang et al. (2016) [56]. This design choice is underpinned by its proven efficacy in classifying time-series data. Unlike recurrent neural networks, which involve extra costs due to sequential processing, the convolutional approach focuses on detecting local temporal features such as the 11–16 Hz oscillatory pattern of sleep spindles (SS) or the biphasic waveform of K-complexes (KC), both of which are essential for the precise identification of sleep events. This approach also draws inspiration from Shelhamer et al. (2017) [62], who demonstrated that fully convolutional networks (FCNs) achieve semantic segmentation from “pixel to pixel” in computer vision; here, it is adapted to execute “time point-to-event probability” mapping in EEG, a key function for real-time decoding of sleep events. The input setup of the backbone is characterized as “batch size × 1 × 300,” wherein “1” denotes the single-channel EEG and “300” represents 3 seconds of EEG data sampled at 100 Hz. This configuration is tailored to encapsulate the full duration of most sleep events while maintaining temporal resolution. Further details on the backbone parameters can be found in the Supplementary materials.

#### 4.5.2 Sliding window strategy

To address the limitation of fixed-epoch analysis in which sleep events may span multiple 3-second intervals and thus be split or missed, a sliding window technique was integrated with the backbone, as shown in Figure 1b. This method converts 3-second chunks of EEG data into continuous event probability distributions, sampled at 100 Hz, encapsulating the concept of “sleep semantic segmentation.” The sliding window matches the input size of the backbone (3 seconds, 300 time points) and moves across the entire EEG record from the night at 10-millisecond steps, corresponding to the EEG’s sampling rate, thereby ensuring temporal continuity in the output. A softmax layer within the system generates a probability vector for each time point, ensuring the total sum is 1 across the seven recognized sleep event categories. The event probability at any given EEG time point is assessed by the outputs from sliding windows centered on that time point.

#### 4.5.3 Model training

The SSSM training was tailored to emphasize robustness against variations in sleep events and ensure generalizability across different populations, which are essential for applying the model to real-world sleep analysis. Each phase was based on the characteristics of the extensive sleep event dataset *D* and the fundamental framework of the SSSM, as detailed below:

Initially, the 3-block convolutional backbone was initialized with weights obtained from the SSL pre-training step. This strategic decision was made to capitalize on EEG features relevant to sleep that were learned from unlabeled data. To prepare the samples, we standardized the input segments to accommodate the variable durations of sleep events in *D*, so they matched the 3-second (300 time points, 100 Hz) input dimension of the backbone. For every annotated event in *D*, we determined its midpoint (for instance, a 1-second spindle occurring between 10 and 11 seconds in the EEG had a midpoint at 10.5 seconds) and extracted a ±1.5-second window centered on this midpoint. This chosen window size ensured comprehensive coverage of even the longest target events (3 seconds) and prevented the inclusion of unrelated adjacent activities. This standardization was crucial as it aligned with the backbone’s receptive field, which is designed to capture 3 seconds of EEG data, thereby permitting the model to consistently extract features across all event types, irrespective of their initial duration. For events like BG, MA, and SW, which exceed 3 seconds, we randomly selected non-overlapping 3-second segments from EEG intervals. Further specifics regarding backbone parameters, including the size of the convolution kernel and the learning rate, can be found in the Supplementary materials.

#### 4.5.4 Computational Efficiency Validation

To ensure the SSSM is applicable in practical scenarios (such as clinical sleep assessment and closed-loop interventions), it is essential that it runs efficiently on conventional hardware. To assess this efficiency, we tested the SSSM on two distinct systems: a CPU (11th Gen Intel® Core™ i7-11800H, 2.30 GHz) and a GPU (NVIDIA GeForce RTX 3060 Laptop GPU). The experimental dataset contained signals representing an hour of sampling at a frequency of 100 Hz, yielding a total of 360,000 data points. For each system configuration, seven separate trials were performed, each consisting of 10 execution cycles, totaling 70 iterations per platform. The mean and standard deviation of processing time were determined across these iterations.

### 4.6 Downstream application configuration

#### 4.6.1 Forecasting imminent sleep events

Forecasting upcoming sleep events is an essential expansion of the Sleep Semantic Segmentation Model (SSSM)’s functionality, facilitating real-time, closed-loop interventions in sleep (such as spindle-targeted memory reactivation or enhancement of slow-wave activity) and mechanistic studies of the dynamics of sleep events. This endeavor utilizes the continuous, 100 Hz event probability outputs from the SSSM to capture temporal dependencies among events, with a process structured to balance physiological validity, model flexibility, and thorough validation.

##### Forecasting Model Design with iTransformer

Predicting upcoming sleep event probabilities from historical SSSM outputs is inherently a multivariate time series forecasting problem: the input is a sequence of 7-dimensional probability vectors (one vector per 10 ms time point, corresponding to the seven event types), and the output is a 7-dimensional vector of predicted probabilities for future time points. To tackle this issue, we chose the iTransformer [63], an enhanced version of the Transformer specifically designed for multivariate time series, as the temporal context module, which is appended to the SSSM’s softmax layer (Figure 1d). The iTransformer was chosen over traditional Transformers or recurrent models (e.g., LSTMs) for two critical reasons. First, it rearranges the attention and feed-forward layers across opposite dimensions: in contrast to conventional Transformers that integrate information across variables (event types) during the attention phase, the iTransformer processes each variable separately before aggregation, thereby preventing artificial cross-contamination of signals specific to each event. This aligns with the SSSM’s design, which treats each event as a distinct but co-occurring signal. Second, the iTransformer has demonstrated state-of-the-art performance on real-world multivariate datasets (e.g., physiological signals, environmental sensors), confirming its ability to capture the noisy, non-stationary temporal patterns inherent in sleep EEG. For model configuration, we defined the input as the preceding 25 seconds of SSSM probability sequences (2,500 time points, 100 Hz) and the output as the subsequent 2 seconds of predicted probabilities (200 time points). This window size was optimized to balance two needs: (1) the 25-second input captures enough historical context to model event dependencies (e.g., the clustering of spindles every 5–10 seconds), and (2) the 2-second output encompasses the typical onset-to-peak duration of most sleep events (0.5–3 seconds), which is essential for interventions that need prior notification (e.g., initiating a stimulus 500 ms earlier than a forecasted SS). We trained and validated the iTransformer-SSSM pipeline exclusively on the MASS-C3 subset of MASS.

MASS-C3 was selected for its demographic diversity (42.5 ± 18.9 years, 55% female) and high-quality EEG recordings, ensuring the model learned generalizable temporal patterns rather than dataset-specific artifacts.

##### Validating Temporal Relationships Between Key Events

To ensure the forecasting model captured biologically meaningful temporal associations (not just statistical noise), we designed two targeted experiments focusing on four clinically relevant events: SS, SW, KC, and MA. These experiments quantified how two factors (past event occurrence count and temporal proximity) influence future event likelihood, with results validated against known sleep physiology. Experiment 1, Influence of Past Event Occurrence Count: We synthesized datasets where the number of past occurrences of a target event (e.g., SS) was systematically varied (2, 3, 4, 5, or 6 occurrences) while controlling for event interval (fixed at 5 seconds between consecutive events) and event tail (minimized to *<*0.1 seconds, ensuring no overlap between events). This control ensured that any changes in predicted future probability were driven solely by occurrence count, not interval or residual activity. For each synthesized dataset, we input the past 25 seconds of probability sequences into the forecasting model and extracted the predicted probability of the target event in the next 2 seconds. We then computed the Pearson correlation coefficient between occurrence count and predicted probability. Experiment 2, Influence of Temporal Proximity to Past Events: We synthesized datasets where the timing of a single past event was adjusted in 0.1-second increments (from 0 to 22 seconds before the forecast window), while again minimizing event tail to avoid residual activity. This design tested how the recency of a past event affects future predictions.

#### 4.6.2 Automatic sleep staging

Automatic sleep staging assigning is a clinically foundational task that also serves as a critical test of the SSSM’s versatility: it requires translating the model’s finegrained event-level representations into coarser, functionally meaningful brain states. Our approach leverages SSSM’s pre-learned sleep event features to avoid redefining task-specific inputs, while integrating temporal context (via Transformer encoders) to capture the sequential nature of sleep stages. Below is a detailed breakdown of the model’s architecture, underlying rationale, and thorough evaluation, all rooted in sleep physiology and aligned with demands for clinical applicability.

##### Single-Epoch Sleep Staging Model: Validating Event-Level Feature Utility

Initially, we created a single-epoch model to verify that the event-centric features of SSSM can directly facilitate sleep staging. This model serves as a “baseline” to validate that SSSM’s backbone captures EEG patterns relevant to stage classification, without relying on cross-epoch context. The input design of the model is intricately linked to the central architecture of SSSM: each 30-second EEG epoch, which serves as the standard unit for sleep staging, was divided into 10 distinct 3-second segments. These segments align with the 3-second input requirement of SSSM’s convolutional backbone, involving a 100 Hz sampling rate and comprising 300 time points per segment. This partitioning ensures we reuse the backbone’s pre-trained event-extraction capabilities (e.g., detecting N2-specific spindles, N3-specific slow waves) without modifying its structure. Each 3-second segment was fed into the SSSM backbone, which output a feature vector encoding event-related patterns. These 10 feature vectors (one for each segment) were then integrated through average temporal pooling. This approach maintains the event distribution throughout the epoch (for instance, an N3 epoch with 20 seconds of slow waves will exhibit greater N3 weights than an N2) while minimizing the impact of transient noise. The pooled vector was finally fed into a linear classifier, which mapped event features to one of five sleep stages. This minimalist design was purposefully employed: the utilization of a linear classifier allowed us to isolate the backbone features of SSSM.

##### Multi-Epoch Sleep Staging Model (SSSM-T): Integrating Temporal Context

Sleep staging is not just a per-epoch task: stages transition sequentially (e.g., N1 → N2 → N3, rarely N3 → Wake directly), and contextual information from adjacent epochs improves classification accuracy, especially for ambiguous stages like N1 (which often lacks distinct events). To capture this sequentiality, we extended the single-epoch model with a Transformer encoder [64], creating the multi-epoch model SSSM-T (Figure 1c). The SSSM-T input was defined as a sequence of 20 consecutive epochs. Each of these 20 epochs was processed identically to the single-epoch model: partitioned into 10×3-second segments, fed into the SSSM backbone, and pooled into a epoch-level feature vector. This yielded a sequence of 20 feature vectors (one per epoch), which was fed into a Transformer encoder with 8 attention heads and a model dimension of 512. Critical to the model’s efficiency and robustness, the SSSM backbone remained unchanged during Transformer training, while only the layers of the Transformer were subjected to fine-tuning. This choice preserved the backbone’s pre-learned event features while optimizing the Transformer to model epoch-to-epoch transitions. The Transformer’s self-attention mechanism allowed it to weigh the importance of adjacent epochs: for example, when classifying an ambiguous N1 epoch, it might prioritize features from the preceding Wake epoch and subsequent N2 epoch.

##### Clinical-Grade Evaluation: Ensuring Generalizability Across Diverse Cohorts

To be of clinical utility, a sleep staging model should consistently perform across diverse populations that differ in demographics, recording methods, and clinical scenarios, thereby avoiding overfitting to limited datasets. We therefore conducted a large-scale evaluation using 14 publicly available Polysomnographic (PSG) datasets (10,155 whole-night recordings total, Further details on these datasets are provided in the Supplementary materials), spanning three decades of data collection, geographically dispersed clinical sites, and diverse groups (healthy adults, elderly individuals, patients with sleep apnea or REM behavior disorder). To mimic real-world clinical deployment, we split the 14 datasets into two groups with strict separation. Training/Internal Testing Group (10 datasets): This included datasets like MASS, Sleep-EDF, and SHHS. For each dataset, we used a per-subject split (75% training, 25% testing)—ensuring no individual subject’s data appeared in both training and testing sets (critical for avoiding overfitting, as a subject’s sleep patterns are consistent across epochs). All training data were used to refine SSSM-T into a clinical-ready variant (SSSM-B). Independent Evaluation Group (4 datasets): This included the 2018 PhysioNet/CinC Challenge dataset, Haaglanden Medisch Centrum (HMC) dataset, and two Dream Open Datasets (DOD-H/O). Crucially, no data from these four datasets were used in any step of SSSM-B’s development. This design allowed us to assess “zero-shot” performance: how the model would perform when deployed to a new clinical site with unseen data. All datasets utilized are publicly available (with necessary approval for those with restricted access), ensuring transparency and reproducibility, which are essential requirements for implementing the model in clinical practice.

#### 4.6.3 Sleep related disease diagnosis

The use of sleep EEG to distinguish sleep-related disorders, like REM behavior disorder (RBD), sleep apnea syndrome, and depression-related sleep disturbances, presents a unique technical challenge: it necessitates the examination of lengthy, continuous EEG recordings (spanning 8–10 hours overnight, or about ^~^10,000 3-second segments) while concurrently identifying two crucial feature types: (1) local pathological markers and (2) extended global patterns. Conventional sequence models face difficulties in balancing these requirements; Long Short-Term Memory networks (LSTMs) struggle with effectively capturing long-range dependencies, whereas standard Transformers [64], although proficient in sequence modeling, encounter *O*(*N* ^2^) computational complexity, rendering them unsuitable for full-night EEG analysis. To solve this issue, we combined the SSSM with Mamba [65], which is a structured state space sequence model optimized for lengthy sequences, forming a framework for disease diagnosis that achieves a balance among efficiency, feature detection, and clinical importance.

##### Mamba for Sleep Disease Diagnosis

Mamba’s architecture is specifically designed to address the shortcomings of traditional models, making it highly suitable for the overnight evaluation of EEGs, which is essential for diagnosing sleep disorders. It boasts features finely tuned to the needs of sleep-related diagnostics. Selective Mechanism: Mamba differs from state space models that utilize fixed transition parameters by employing input-dependent parameters, enabling it to dynamically focus on clinically significant EEG segments (such as REM phases in patients with RBD and clusters of arousal induced by apnea during sleep apnea) while excluding irrelevant noise (like stable background activity in healthy N2 sleep). Scan Module: By analyzing the EEG sequence in overlapping windows, the scan module can detect both local pathological markers (such as a 3-second micro-arousal) and long-range dependencies. This method mirrors the diagnostic approach clinicians use for sleep disorders, which considers both transient events and the entire sleep pattern. Linear Computational Complexity: The hardware-optimized algorithm of Mamba ensures linear scalability with sequence length (*O*(*N* )), contrasting with Transformers, which scale quadratically. This characteristic allows Mamba to process efficiently the complete set of 3-second EEG segments from an overnight study (up to 15,000 segments).

##### Mamba-SSSM Fusion Model Design

The disease diagnosis framework leverages the event-level feature extraction capabilities of the SSSM, incorporating a Mamba-based temporal context module to capture long-range EEG dynamics (Figure 1e). This approach allows us to utilize the SSSM’s pre-established sleep event features while introducing the capacity to associate these events with overarching disease patterns. The procedure is organized as follows. Step 1, Generating Disease-Relevant Token Features: Initially, we transform the complete overnight EEG into a sequence of event-level tokens via the SSSM backbone, maintaining the alignment with the core architecture of the model (as elaborated in “Sleep Semantic Segmentation Model Design”). For each 3-second EEG segment (corresponding to the backbone’s input size), the SSSM backbone derives a token that encapsulates event patterns. This token generation phase is vital: it translates raw EEG data into a succinct representation of sleep events, which serve as the biological substrates of disease-associated sleep phenotypes. Step 2, Mamba for Long-Sequence Classification: Subsequently, these tokens are input into a Mamba-based temporal context module, which learns to translate the sequence of event tokens into a disease label. More formally, the process is characterized as:

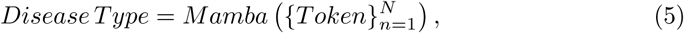

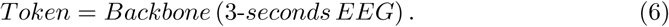

In this context, *N* refers to the number of 3-second intervals in an overnight recording, limited to a maximum of 15,000 (approximately 12.5 hours of EEG, which is more than sufficient for standard 8-10 hour sleep studies). For shorter recordings, such as a 6-hour EEG resulting in about 7,200 segments, we extend the token sequence with zeros to reach 15,000. This approach ensures a consistent input size across different subjects, thus preventing any bias towards longer recordings and facilitating batch training. Importantly, during Mamba training, the SSSM backbone remains unchanged, allowing only the Mamba module to be fine-tuned. This strategy maintains the backbone’s capacity to consistently extract event features while allowing the Mamba module to adapt and learn patterns specific to the disease.

##### Clinical Validation

To ensure the model’s performance reflects real-world clinical utility, we evaluated it on three representative sleep disorders using two publicly available, clinically curated datasets: the CAP Sleep Database and the Sleep Heart Health Study (SHHS). Each dataset was selected for its ability to test the model’s ability to distinguish distinct pathological phenotypes, with strict sample selection criteria aligned with clinical standards. REM Behavior Disorder (RBD) –We used the CAP dataset, which includes 22 overnight recordings from patients with clinically diagnosed RBD and 16 recordings from age-matched healthy controls (no neurological or sleep disorders). Sleep Apnea Syndrome –We selected a subset of SHHS (a large communitybased cohort) with clear clinical stratification. Apnea Group: 262 subjects with an Apnea-Hypopnea Index (AHI) *>* 50 events/hour (severe sleep apnea, per AASM clinical criteria). Healthy Control Group: 215 subjects with both obstructive and central AHI *<* 5 events/hour (no clinically significant sleep apnea). This large sample size (477 total subjects) ensures the model’s performance is not biased by small-sample variability, while the strict AHI thresholds align with how clinicians diagnose sleep apnea. Depression-Related Sleep Disturbance –We used another SHHS subset, where subjects were stratified by a validated “happy index” (a self-reported measure of mood, correlated with depressive symptoms). Depression Group: 91 subjects with a low happy index (score *<* 3/5). Healthy Control Group: 130 subjects with a high happy index (score *>* 4/5)—no self-reported mood disorders or sleep complaints. This stratification leverages SHHS’s comprehensive phenotypic data to ensure the model is not just classifying arbitrary groups, but capturing sleep-related markers of a clinically relevant condition.

## Declarations

### Data availability

The sleep event dataset is openly available on GitHub.

### Code availability

The source code for the SSSM implementation is publicly accessible on the Python Package Index (PyPI) to support reproducibility and ongoing development. Additionally, a web showcase is available at Hugging Face.

