## Supplementary information for "Semantic Segmentation of Sleep Events for High-Resolution Sleep Decoding"

#### This PDF file includes:

Supporting text
Fig. S1
Tables S1 to S5
Legend for Dataset S1
SI References

#### Other supporting materials for this manuscript include the following:

Dataset S1

### Supporting Information Text

**A. Guidelines for Annotating Sleep Events.** Figure S1a-g presents representative raw EEG samples for each of the seven sleep events.

**SS** exhibits a frequency of 11–16 Hz (typically 12–14 Hz) and is most prominent over central regions. These events last at least 0.5 seconds (ranging from 0.5 to 1.5 seconds) and are a hallmark of stage N2 sleep, involving thalamocortical oscillations linked to the reticular thalamic nucleus. They might also be observed during stage N3 sleep. When induced by benzodiazepines, these spindles can have marginally higher frequencies.

**KC** exhibits a prominent high-amplitude biphasic waveform, starting with a distinct negative sharp wave (upwards) followed by a positive slow wave (downwards), clearly emerging from the lower voltage background. The duration is at least 0.5 seconds, and it is a feature of stage N2 sleep, with the strongest signals over the frontal areas (frontal > central > occipital). An arousal associated with a KC must begin within 1 second of the KC's end.

**SW**, with a frequency of 0.5–2 Hz and a peak-to-peak amplitude exceeding 75 $\mu V$ in frontal regions, are used to identify stage N3 sleep. In stage N2, SWA constitutes less than 20% (< 6 seconds), whereas in stage N3, it constitutes at least 20% (6 seconds or more), and SWA is frequently observed in eye derivations.

**SAW**, noted for their train of serrated triangular patterns, range from 2–6 Hz in frequency and peak in amplitude within central regions. These waves often precede a burst of REM, though they are not essential for its identification.

**VSW** stands out with sharply defined contours. Its duration is less than 0.5 seconds, with maximum prominence over the central area (areas including C3, C4, Cz). These waves are distinct from the surrounding activity with higher amplitudes and occur in stage N1, typically near the transition to stage N2, and also manifest during stage N2.

**MA** represents a sudden change in EEG frequency patterns, involving alpha, theta, and/or frequencies above 16 Hz, excluding spindles, that persists for a minimum of 3 seconds, with at least 10 seconds of stable sleep preceding the change. When scoring MA during REM sleep, there must be a simultaneous increase in submental EMG activity lasting at least 1 second. This study also categorized W as MA. W features an EEG baseline of continuous, symmetrical, and irregular mixed frequencies of low to medium amplitude, potentially comprising: a) irregular theta and delta activity (up to 100 $\mu V$) most prominent in O1, O2; b) widespread irregular alpha and beta activity (up to 30 $\mu V$); c) rhythmic theta activity (up to 50 $\mu V$), often most evident in C3, Cz, C4; d) artifacts due to body and eye movements.

**BG** lacks a precise definition in the AASM, yet it consistently appears as a criterion for exclusion when defining the sleep events mentioned above. In this research, BG is categorized as any activity that does not encompass SS, KC, SW, SAW, VSW, or MA.

**B. Statistical information of this sleep event dataset.** We conducted a by-subject count analysis of each event occurrences in MASS dataset, and the scatter plot of these occurrences is shown in Figure S1h. Each subject has 256±147 BGs, 90±105 KCs, 133±77 MAs, 17±29 SAWs, 398±367 SSs, 351±231 SWs, and 8±10 VSWs. Figure S1i presents stratified statistics based on age and gender for three metrics: number of occurrences, duration, and Peak-to-Peak (PTP) amplitude, organized in rows. Seven sleep events are displayed in columns.

Sleep event count differences associated with age (Younger cohort: 100 individuals aged 18–35; Older cohort: 100 individuals aged 50–76) and sex (88 females (F) and 92 males (M)) in MASS were analyzed. A 2 × 2 ANOVA indicated significant effects for age ($F(1,1331)=109.2$, $p<0.001$) and sex ($F(1,1331)=13.46$, $p<0.001$). Age differences in BG counts were significant only for males ($F(1,1331)=15.23$, $p<0.001$). For KC counts, significant age differences were observed in both males ($F(1,1331)=7.07$, $p=0.008$) and females ($F(1,1331)=15.85$, $p<0.001$). SW counts showed significant age differences in both males ($F(1,1331)=116.543$, $p<0.001$) and females ($F(1,1331)=45.67$, $p<0.001$). In the older group, a significant sex difference in SW counts was found ($F(1,1331)=26.18$, $p<0.001$). Significant age differences in SS counts were observed for males ($F(1,1331)=90.78$, $p<0.001$) and females ($F(1,1331)=123.59$, $p<0.001$), with sex differences present in both younger ($F(1,1331)=25.31$, $p<0.001$) and older ($F(1,1331)=9.27$, $p=0.002$) groups.

**C. Data source description.** A total of 17 publicly available datasets were employed in this study. Table S1 provides a brief overview of the details for these datasets.

The **Montreal Archive of Sleep Studies (MASS)** (1) is an open-access and collaborative database of laboratory-based polysomnography (PSG) recordings. This cohort comprises polysomnograms of 200 complete nights recorded in 97 men and 103 women of age varying between 18 and 76 years (mean: 38.3 years, SD: 18.9 years). It has been split in five different subsets: MASS-C1, MASS-C2, MASS-C3, MASS-C4 and MASS-C5. MASS-C2, MASS-C3 and MASS-C5 were scored by 20-seconds epochs. Only MASS-C1 and MASS-C3 were scored by 30-seconds epochs, classifying each segment into one of eight categories: {'W', 'N1', 'N2', 'N3', 'N4', 'REM', and '?'}. Conforming with AASM guidelines, the N3 and N4 classes were combined into a single N3 category, while the '?' classes were excluded. In this study, C3-M2 channel was selected for all analysis.

The **Sleep Heart Health Study (SHHS)** (2, 3) is a thorough, multi-center cohort dataset aimed at exploring the cardiovascular and other health outcomes related to sleep-disordered breathing. It comprises two phases: SHHS1 and SHHS2. In the PSG of each phase, recordings include two bipolar EEG channels (C4-M1, C3-M2), one EKG channel, two EOG channels, two lower limb EMG channels, snoring detection, pulse oximetry, and a body position sensor. Sleep experts analyzed these recordings, segmenting them every 30 seconds into eight predefined categories: 'W', 'N1', 'N2', 'N3', 'N4', 'REM', 'Movement', and 'Unknown'. Conforming to the AASM guidelines, the N3 and N4 categories were consolidated under N3, while 'Movement' and 'Unknown' categories were excluded. In this study, C3-M2 channel was selected for all analysis.

This research employed the expanded **Sleep-EDF dataset (SEDF)** (4), comprising two distinct subsets: SC and ST. The SC subset includes PSG data from 153 healthy individuals ranging from 25 to 101 years old. Conversely, the ST subset consists of PSG data from 44 individuals aged 18 to 79, specifically selected to assess the effects of temazepam on sleep. The PSG configuration features two bipolar EEG channels (Fpz-Cz and Pz-Oz), a horizontal EOG channel, and a submental chin EMG channel. Sleep staging was conducted every 30 seconds by sleep specialists, classifying each segment into one of eight categories: {'W', 'N1', 'N2', 'N3', 'N4', 'REM', 'M', and '?'}. Conforming with AASM guidelines, the N3 and N4 classes were combined into a single N3 category, while the 'M' and '?' classes were excluded. In this study, Fpz-Cz channel was selected for all analysis.

The **ISRUC-Sleep** dataset (5) is composed of three subsets aimed at examining both individuals without sleep disorders and those on sleep medication. The first subset contains data from 100 subjects, each having a single PSG recording. The second subset comprises data from 8 individuals, each undergoing two PSG recordings. The third subset involves data from 10 healthy subjects, each with one PSG recording. Within this dataset, two sleep specialists annotated each 30-second segment with one of the following five classifications: 'W', 'N1', 'N2', 'N3', 'REM'. This research utilized the EEG signal from C3-M2 channel and utilized the annotations from the first sleep specialist.

The **CAP** Sleep Database (6) is a collection of 108 PSG recordings. The recordings include at least 3 EEG channels (F3 or F4, C3 or C4 and O1 or O2, referred to M1 or M2), EOG (2 channels), EMG of the submentalis muscle, bilateral anterior tibial EMG, respiration signals (airflow, abdominal and thoracic effort and SaO2) and EKG. The 16 healthy subjects included in the study did not present any neurological disorders and were free of drugs affecting the central nervous system. The 92 pathological recordings include 40 recordings of patients diagnosed with Nocturnal frontal lobe epilepsy, 22 affected by REM behavior disorder, 10 with Periodic leg movements, 9 insomniac, 5 narcoleptic, 4 affected by Sleep-disordered breathing and 2 by bruxism.Nonetheless, these recordings display some inconsistency in the channels, with certain recordings lacking the C3-M2 channel while possessing the C4-M1, and vice versa. Consequently, this study consistently selected either the C4-M1 or C3-M2 channel for all analyses. Sleep staging was conducted every 30 seconds by sleep specialists, classifying each segment into one of eight categories: {'W', 'N1', 'N2', 'N3', 'N4', 'REM' and 'MT'}. Conforming with AASM guidelines, the N3 and N4 classes were combined into a single N3 category, while the 'MT' class were excluded.

**MROS** (7, 8) is an ancillary study of the parent Osteoporotic Fractures in Men Study. Between 2000 and 2002, 5,994 community-dwelling men 65 years or older were enrolled at 6 clinical centers in a baseline examination. Between December 2003 and March 2005, 3,135 of these participants were recruited to the Sleep Study when they underwent full unattended PSG and 3 to 5-day actigraphy studies. The objectives of the Sleep Study are to understand the relationship between sleep disorders and falls, fractures, mortality, and vascular disease. Sleep staging was conducted every 30 seconds by sleep specialists, classifying each segment into one of six categories: {'W', 'N1', 'N2', 'N3', 'N4' and 'REM'}. Conforming with AASM guidelines, the N3 and N4 classes were combined into a single N3 category. In this study, C3-M2 channel was selected for all analysis.

**Haaglanden Medisch Centrum (HMC)** (9) sleep staging database A collection of 151 whole-night polysomnographic (PSG) sleep recordings (85 Male, 66 Female, mean Age of 53.9 $\pm$ 15.4) collected during 2018 at the Haaglanden Medisch Centrum (HMC, The Netherlands) sleep center. The PSG data consist of four EEG (F4-M1, C4-M1, O2-M1, and C3-M2), two EOG (E1-M2 and E2-M2), one bipolar chin EMG, and one ECG (single modified lead II) derivations. Current guidelines for sleep scoring carry out segmentation of neurophysiological activity in discrete 30s epochs. Each epoch may be classified as one of five possible states according to signal activity: {W, stages N1, N2, N3, and 'REM'}. In this study, C3-M2 channel was selected for all analysis.

The **Dreem** Open Dataset (10) – Healthy (DOD-H) was collected from 25 volunteers at the French Armed Forces Biomedical Research Institute's Fatigue and Vigilance Unit in France. Subjects were without sleep complaints, aged 18-65 and locally recruited without regard to gender or ethnicity. The Dreem Open Dataset Obstructive (DOD-O) was collected from the Stanford Sleep Medicine Center, California, US from 55 patients (clinical trial NCT03657329) with clinical suspicion for sleep-related breathing disorder. Individuals clinically diagnosed with sleep disorders other than OSA, suffering from morbid obesity, taking sleep medications or with certain cardiopulmonary or neurological comorbidities were exclude from the study. Sleep staging was conducted every 30 seconds by sleep specialists, classifying each segment into one of seven categories: {'W', 'N1', 'N2', 'N3', 'N4' , 'REM' and 'NOT SCORED'}. In this study, the 'NOT SCORED' class were excluded, and C3-M2 channel was selected for all analysis.

The over-night PSG data the from **2018 PhysioNet/CinC Challenge (Challenge2018)** (11) were contributed by the Massachusetts General Hospital's Computational Clinical Neurophysiology Laboratory and the Clinical Data Animation Laboratory. The full dataset spans 1,985 patients who were monitored for the diagnosis of sleep disorders. The original challenge was automatic detection of arousal, but sleep stages were annotated by clinical staff. Sleep staging was conducted every 30 seconds by sleep specialists, classifying each segment into one of six categories: {'W', 'N1', 'N2', 'N3', and 'REM'}. In this study, C3-M2 channel was selected for all analysis.

**D. Training settings and hyperparameters.** The model's training and evaluation were performed on a computer featuring an Intel I9-12900K CPU running at 5.20GHz, 64GB of RAM, and an NVIDIA GPU 3090. Additionally, all data processing and algorithm development was executed using Python version 3.11.5, alongside the Pytorch library version 2.2.0. The number of training epochs was fixed at 30 for all experimental trials. The fundamental hyperparameters are outlined as follows. Batch size was designated as 32 across the experiments involving the backbone, sleep stage (single epoch), sleep stage (multiple epochs), and sleep event forecasting. For the sleep-related disease diagnosis model training, the batch size was set to 10. The learning rate was fixed at 1.00E-04 for the backbone, sleep event forecasting, and sleep-related disease diagnosis model training. It was adjusted to 2.00E-06 and 3.00E-05 for training the sleep stage (single epoch) and sleep stage (multiple epochs) models, respectively. The convolution block shapes for the SSSM are (1, 32) for block 1, (32, 64) for block 2, and (64, 128) for block 3. The fully connected layer's shape is (15×128, 7). The downstream hyperparameters are as follows. During the training of the sleep stage (single epoch) model, the SSSM sample overlap was set to 0, with a sequence length of 10, 8 encoder layers, 32 attention heads, and a model dimension of 1024. For the sleep stage (multiple epochs) model, the SSSM sample overlap was 0, with a sequence length of 200, 1 encoder layer, 32 attention heads, and a model dimension of 512. In the sleep event forecasting model, the SSSM sample overlap was 299, with 2 encoder layers, 32 attention heads, and a model dimension of 128. For the sleep-related disease diagnosis model training, the SSSM sample overlap was 0, a model dimension of 64, an SSM state expansion factor of 64, a local convolution width of 32, and a block expansion factor of 4.

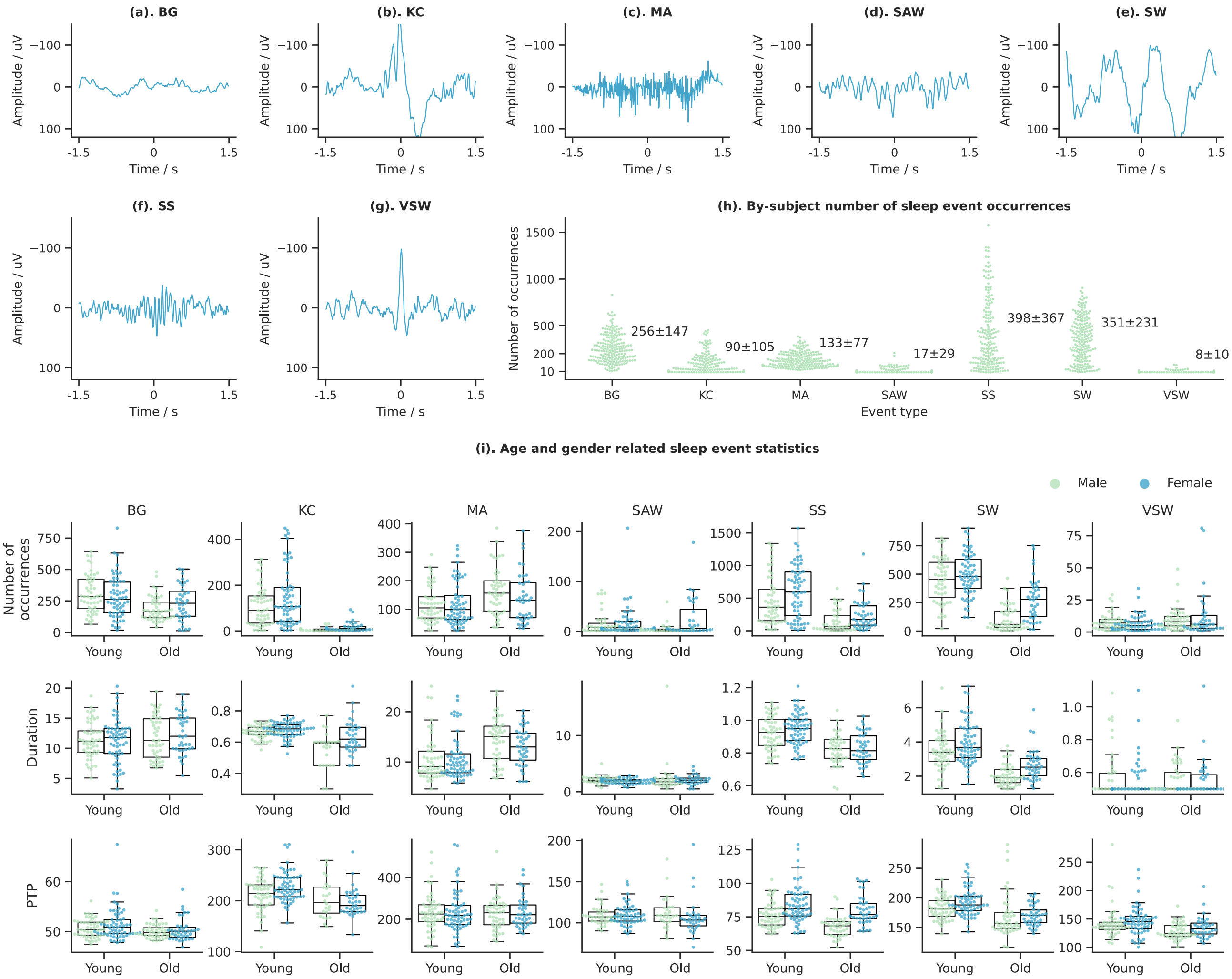


**Fig. S1.** **(a)-(g).** Representative Sleep event samples. This illustration presents a prototypical 3-second data window for various sleep events: Background (BG), K-complex (KC), Micro-arousal (MA), Sawtooth wave (SAW), Sleep spindles (SS), Slow waves (SW), and Vertex sharp waves (VSW). **(h).** Scatter plots of sleep event occurrences. The X-axis labels the different sleep events. The X-axis denotes distinct sleep events, while the Y-axis displays the number of occurrences. Adjacent to each event's scatter plot are the mean and standard deviation (mean±std). **(i).** Stratified sleep event statistics based on age and gender. Stratified analyses of age and gender were conducted on three metrics: number of occurrences, duration, and Peak-to-peak (PTP) amplitude, which are organized in rows, and the seven sleep events are arrayed in columns.

| Dataset | Records | Age (years) | Sex % (F/M) | Channel selected |
|---|---|---|---|---|
| MASS-C1 | 53 | 63 ± 5.3 | 36/64 | C3-M2 |
| MASS-C2 | 19 | 23.6 ± 3.7 | 58/42 | C3-M2 |
| MASS-C3 | 62 | 42.5 ± 18.9 | 55/45 | C3-M2 |
| MASS-C4 | 40 | 25.3 ± 4.3 | 65/35 | C3-M2 |
| MASS-C5 | 26 | 25 ± 7.4 | 50/50 | C3-M2 |
| SEDF-SC | 153 | 58.8 ± 22.0 | 53/47 | Fpz-Cz |
| SEDF-ST | 44 | 40.2 ± 17.7 | 68/32 | Fpz-Cz |
| ISRUC-SG1 | 100 | 51.1 ± 15.9 | 44/56 | C3-M2 |
| ISRUC-SG2 | 16 | 46.9 ± 17.5 | 25/75 | C3-M2 |
| ISRUC-SG3 | 10 | 39.6 ± 9.6 | 10/90 | C3-M2 |
| SHHS | 8444 | 63.1 ± 11.2 | 52/48 | C3-M2 |
| CAP | 108 | 45.2 ± 19.6 | 76/24 | C4-M1 or C3-M2 |
| MROS | 3926 | 76.4 ± 5.5 | 0/100 | C3-M2 |
| HMC | 151 | 53.9 ± 15.4 | 44/56 | C3-M2 |
| DOD-H | 25 | 35.3 ± 7.5 | 24/76 | C3-M2 |
| DOD-O | 55 | 45.6 ± 16.5 | 36/64 | C3-M2 |
| Challenge2018 | 1985 | 55 ± 14.4 | 34/65 | C3-M2 |

**Table S1. Details of Sleep Datasets Used in the Study**

**Table S2. Comparison with other methodologies about linear evaluation using single-epoch EEG.**

| | Sleep-EDFX | | | | | | | SHHS | | | | | | | ISRUC-Sleep | | | | | | |
|---|---|---|---|---|---|---|---|---|---|---|---|---|---|---|---|---|---|---|---|---|---|
| | Per-Class F1 | | | | | Overall | | Per-Class F1 | | | | | Overall | | Per-Class F1 | | | | | Overall | |
| | W | N1 | N2 | N3 | REM | ACC | MF1 | W | N1 | N2 | N3 | REM | ACC | MF1 | W | N1 | N2 | N3 | REM | ACC | MF1 |
| SimCLR | 85.95 | 32.25 | 79.44 | 68.65 | 53.59 | 71.49 | 63.98 | 83.61 | 23.14 | 80.64 | 82.75 | 71.92 | 77.75 | 68.41 | 82.24 | 39.41 | 69.38 | 78.43 | 68.31 | 70.98 | 67.55 |
| BYOL | 87.08 | 33.40 | 78.32 | 64.49 | 55.12 | 71.82 | 63.68 | 83.20 | 19.50 | 81.71 | 84.52 | 71.32 | 78.51 | 68.05 | 82.30 | 40.62 | 70.42 | 77.48 | 67.76 | 71.22 | 67.72 |
| SwAV | 85.11 | 32.49 | 78.92 | 62.68 | 54.44 | 71.98 | 62.73 | 81.88 | 21.80 | 80.10 | 82.17 | 71.93 | 77.21 | 67.58 | 80.22 | 38.50 | 68.00 | 75.60 | 66.15 | 68.91 | 65.69 |
| SimSiam | 85.55 | 29.30 | 78.92 | 63.62 | 46.89 | 71.34 | 60.85 | 84.77 | 21.06 | 81.59 | 83.56 | 73.09 | 79.23 | 68.81 | 81.55 | 38.05 | 68.74 | 76.13 | 66.05 | 69.68 | 66.10 |
| Barlow Twins | 87.85 | 29.20 | **81.84** | 69.69 | 57.33 | 75.13 | 65.18 | 84.36 | 23.84 | 81.69 | 83.82 | 74.13 | 79.27 | 69.57 | 83.84 | 40.02 | 71.24 | 78.65 | 68.68 | 72.23 | 68.49 |
| MAE | 81.23 | 27.39 | 76.72 | 60.99 | 46.91 | 68.31 | 58.65 | 85.28 | 18.38 | 83.95 | 84.90 | 75.86 | 81.08 | 69.67 | 81.94 | 36.41 | 74.01 | 83.69 | 58.42 | 71.49 | 66.89 |
| SimMIM | 83.68 | 26.69 | 75.73 | 51.32 | 44.32 | 69.94 | 56.35 | 84.94 | 22.38 | 83.14 | 85.11 | 75.25 | 80.45 | 70.16 | 82.05 | 35.75 | 74.43 | 84.13 | 58.79 | 71.64 | 67.03 |
| Data2Vec | 83.39 | 28.54 | 75.41 | 56.54 | 50.82 | 69.78 | 58.94 | 78.07 | 16.65 | 79.08 | 81.02 | 71.24 | 76.18 | 65.21 | 82.22 | 36.59 | 73.42 | 83.09 | 60.83 | 71.71 | 67.23 |
| BENDR | 72.15 | 28.28 | 67.83 | 50.94 | 32.5 | 57.42 | 50.34 | 52.34 | 08.41 | 72.72 | 78.37 | 54.78 | 65.08 | 53.32 | 52.75 | 13.83 | 72.18 | 78.28 | 55.69 | 64.78 | 54.55 |
| ContraWR | 88.40 | 34.35 | 81.67 | 68.81 | 62.78 | 75.79 | 67.20 | 85.78 | **25.51** | 84.20 | 85.79 | 77.53 | 81.65 | 71.76 | 84.09 | 40.23 | 73.26 | 82.55 | **71.18** | 74.07 | 70.26 |
| TS-TCC | 73.28 | 21.15 | 66.01 | 41.39 | 37.33 | 61.45 | 47.83 | 70.05 | 17.68 | 75.33 | 73.23 | 62.00 | 70.43 | 59.66 | 80.91 | 32.06 | 70.13 | 80.27 | 64.17 | 70.17 | 65.51 |
| mulEEG | 89.09 | 36.52 | 80.69 | 69.62 | 59.66 | 74.92 | 67.12 | 83.67 | 21.41 | 83.06 | 85.82 | 74.16 | 79.94 | 69.62 | 80.87 | 36.69 | 71.25 | 82.56 | 65.77 | 71.58 | 67.43 |
| NeuroNet-T | **89.90** | 30.21 | 81.51 | **71.39** | 59.51 | **76.26** | 66.50 | 83.97 | 13.95 | 83.30 | 85.45 | 73.75 | 80.45 | 68.09 | 84.51 | 39.40 | **76.15** | **84.68** | 67.63 | **75.12** | **70.47** |
| NeuroNet-B | **89.17** | **36.24** | **81.74** | 69.97 | **63.82** | **76.74** | **68.19** | **88.27** | **30.76** | **86.20** | **87.56** | **79.41** | **84.13** | **74.44** | **85.08** | **42.11** | **76.84** | **85.74** | **71.04** | **76.47** | **72.16** |
| SSSM | 88.94 | **32.59** | 80.56 | 77.39 | **62.42** | 75.23 | 68.38 | **87.73** | 11.79 | **85.41** | **86.70** | **77.89** | **83.30** | 69.90 | **84.91** | 42.79 | 73.98 | 82.80 | 65.52 | 73.77 | 70.05 |

The highest value in each column is highlighted with an underline and bold text, the second highest is highlighted in bold, and the third highest is underlined.

**Table S3. Comparision between supervised learning-based methodologies and SSSM+T.**

| | Sleep-EDFX | | | | | | | SHHS | | | | | | | ISRUC-Sleep | | | | | | |
|---|---|---|---|---|---|---|---|---|---|---|---|---|---|---|---|---|---|---|---|---|---|
| | Per-Class F1 | | | | | Overall | | Per-Class F1 | | | | | Overall | | Per-Class F1 | | | | | Overall | |
| | W | N1 | N2 | N3 | REM | ACC | MF1 | W | N1 | N2 | N3 | REM | ACC | MF1 | W | N1 | N2 | N3 | REM | ACC | MF1 |
| DeepSleepNet | 90.84 | 35.56 | 81.42 | 68.59 | 68.02 | 77.49 | 68.89 | 83.84 | 18.87 | 83.55 | 84.67 | 76.8 | 81.02 | 69.55 | 81.55 | 38.25 | 68.9 | 81.17 | 62.17 | 69.84 | 66.41 |
| IITNet | 70.05 | 45.77 | 83.61 | 63.65 | 80.9 | 81.48 | 73.3 | 89.32 | 47.38 | 85.57 | 80.96 | 87.58 | 84.74 | 78.16 | 84.6 | 40.51 | **78.39** | 85.27 | **79.15** | **77.89** | 73.59 |
| U-Sleep | 92.71 | 47.72 | 84.65 | 65.23 | **82.44** | 82.42 | 74.55 | 89.56 | 48.08 | 86.53 | 81.76 | **88.82** | 85.59 | 78.95 | **86.34** | 44.16 | **79.07** | 85.38 | **81.48** | **78.89** | 75.29 |
| AttnSleep | 91.49 | 40.44 | 83.84 | 72.28 | 72.18 | 79.68 | 72.05 | 87.28 | 28.13 | 83.83 | 85.39 | 77.72 | 81.98 | 72.47 | 84.54 | 42.41 | 75.81 | 83.45 | 69.92 | 75.72 | 71.23 |
| SleepExpertNet | 92.8 | 52.75 | **85.92** | 73.4 | 80.93 | **83.13** | **77.16** | **90.84** | 40.71 | 86.60 | 84.04 | 87.94 | 85.94 | 78.03 | 72.59 | 14.25 | 66.61 | 72.69 | 58.05 | 64.89 | 56.84 |
| NeuroNet-T+TCM | 92.27 | **53.01** | 85.19 | 75.23 | 77.13 | 82.67 | 76.57 | 86.44 | **50.17** | 85.97 | 85.97 | 83.99 | 84.62 | 78.51 | 83.75 | 44.73 | 77.50 | **86.61** | 72.04 | 76.79 | 72.93 |
| NeuroNet-B+TCM | **93.15** | **58.80** | **87.21** | **76.97** | **83.00** | **85.24** | **79.82** | 89.05 | **55.29** | **88.09** | **86.48** | 87.25 | **86.88** | **81.23** | 84.5 | **46.09** | 77.25 | **86.86** | 72.57 | 77.05 | 73.45 |
| SSSM-T | **92.91** | 46.92 | 84.79 | **79.04** | 79.33 | 82.78 | 76.60 | **91.09** | 46.47 | **86.69** | **87.01** | **88.99** | **87.87** | **80.45** | **86.01** | **53.35** | 76.04 | 86.15 | 75.37 | 77.21 | **75.38** |

The highest value in each column is highlighted with an underline and bold text, the second highest is highlighted in bold, and the third highest is underlined.

**Table S4. Overall Performance of SSSM based Big Sleep Stage Model**

| Type | | Per-Class MF1 | | | | | Overall | |
|---|---|---|---|---|---|---|---|---|
| | | W | N1 | N2 | N3 | REM | ACC | MF1 |
| Internal | a | 90.77±5.08 | 44.63±2.56 | 83.21±5.85 | 76.98±6.05 | 86.74±1.87 | 82.71±4.72 | 76.46±2.50 |
| | b | 94.11±1.97 | 45.68±2.74 | 87.95±1.31 | 76.68±8.22 | 87.80±0.85 | 88.45±1.66 | 78.45±2.19 |
| Hold-Out | a | 80.73±2.95 | 42.29±9.23 | 82.23±3.41 | 77.67±3.05 | 80.43±2.36 | 78.19±3.14 | 72.69±2.85 |
| | b | 77.84±2.47 | 36.24±3.25 | 81.70±1.67 | 76.16±1.57 | 81.74±1.22 | 75.68±1.36 | 70.74±1.04 |
| Overall | a | 87.90±6.48 | 43.96±5.04 | 81.70±1.67 | 77.18±5.26 | 84.94±3.53 | 81.42±4.71 | 75.39±3.58 |
| | b | 88.70±7.96 | 42.54±5.32 | 85.87±3.27 | 76.51±6.78 | 85.78±3.02 | 84.20±6.22 | 75.88±4.09 |

[a] not weighted by number of test records.
[b] weighted by number of test records

**Table S5. Performance of sleep related disease diagnosis**

| Disease | Dataset | Per-class records | Per class MF1 | Overall ACC | Overall MF1 |
|---|---|---|---|---|---|
| Sleep apnea syndrome | SHHS | H: 215<br>A: 262 | H: 80.85± 2.77<br>A: 77.30±2.07 | 79.33±1.86 | 79.07±1.79 |
| Depression | SHHS | H: 130, D: 91 | H: 68.84±16.06<br>D: 79.97±9.86 | 75.65±10.91 | 73.40±11.87 |
| REM behavior disorder | CAP | H: 16<br>R: 22 | H: 80.00±18.26<br>R: 88.00±10.95 | 85.00±13.69 | 84.00±14.60 |

[H] Health
[A] Sleep apnea syndrome
[R] REM behavior disorder
[D] Depression

**SI Dataset S1 (Sleep_event_annotations.csv)**

Includes the annotation data for sleep events as presented in Table 1, detailing start time, end time, file name of the sleep recording, and label name.
